# Risk of cancer in regular and low meat-eaters, fish-eaters, and vegetarians: a prospective analysis of UK Biobank participants

**DOI:** 10.1101/2021.09.15.21263656

**Authors:** Cody Z. Watling, Julie A. Schmidt, Yashvee Dunneram, Tammy Y. N. Tong, Rebecca K. Kelly, Anika Knuppel, Ruth C. Travis, Timothy J. Key, Aurora Perez-Cornago

**Affiliations:** Cancer Epidemiology Unit, Nuffield Department of Population Health, University of Oxford, Oxford, United Kingdom

**Keywords:** Diet, vegetarian, pescatarian, cancer, prospective, meat

## Abstract

**Background:** Following a vegetarian diet has become increasingly popular and some evidence suggests that being vegetarian may be associated with a lower risk of cancer overall. However, for specific cancer sites, the evidence is limited.

**Aim:** To assess the associations of vegetarian and non-vegetarian diets with risks of all cancer, colorectal cancer, postmenopausal breast cancer, and prostate cancer, and to explore the role of potential mediators between these associations.

**Methods:** We conducted a prospective analysis of 472,377 UK Biobank participants who were free from cancer at recruitment. Participants were categorised into regular meat-eaters (n=247,571), low meat-eaters (n=205,385), fish-eaters (n=10,696), and vegetarians (n=8,685) based on dietary questions completed at recruitment. Multivariable-adjusted Cox regressions were used to estimate hazard ratios (HR) and 95% confidence intervals (CI) for all cancer incidence and separate cancer sites across diet groups.

**Results:** After an average follow-up of 11.4 years, 54,961 incident cancers were identified, including 5,882 colorectal, 7,537 postmenopausal breast, 9,501 prostate cancer cases. Compared with regular meat-eaters, being a low meat-eater, fish-eater, or vegetarian were all associated with a lower risk of all cancer (HR: 0.98, 95% CI: 0.96-1.00; 0.90, 0.84-0.96; 0.86, 0.80-0.93, respectively). Being a low meat-eater was associated with a lower risk of colorectal cancer in comparison to regular meat-eaters (0.91, 0.86-0.96); there was heterogeneity in this association by sex (p=0.007), with an inverse association across diet groups in men, but not in women. Vegetarian postmenopausal women had a lower risk of breast cancer (0.82, 0.68-0.99), which was attenuated and non-significant after adjusting for body mass index (BMI; 0.87, 0.72-1.05); in mediation analyses, BMI was found to possibly mediate the observed association. In men, being a fish-eater or a vegetarian was inversely associated with prostate cancer risk (0.80, 0.65-0.99 and 0.69, 0.54-0.89, respectively).

**Conclusion:** Low and non-meat-eaters had a lower risk of being diagnosed with cancer in comparison to regular meat-eaters. We also found that low meat-eaters had a lower risk of colorectal cancer, vegetarian women had a lower risk of postmenopausal breast cancer, and vegetarians and fish-eaters had a lower risk of being diagnosed with prostate cancer. The lower risk of colorectal cancer in low meat-eaters is consistent with previous evidence suggesting an adverse impact of meat intake. The lower risk of postmenopausal breast cancer in vegetarian women may be explained by their lower BMI. It is not clear whether the other differences observed, for all cancers and for prostate cancer, reflect any causal relationships or are or due to other factors such as residual confounding or differences in cancer detection.

## Introduction

Cancer is a leading cause of death worldwide^1^, and in the United Kingdom (UK) 28% of all deaths in 2017 were attributable to a cancer diagnosis^2-4^. Colorectal, breast, and prostate cancer collectively account for 39% of all new cancer diagnoses in the UK^5^, and it has been estimated that nearly 40% of cancer diagnoses may be preventable through modifiable factors^6, 7^. Although several dietary factors have been suggested to influence cancer risk, it remains unclear whether dietary patterns are related to the risk of developing cancer^8^.

It has been hypothesized that vegetarian diets, which exclude the consumption of all meat and fish, may be associated with a lower cancer risk. In addition to excluding red and processed meat, which are associated with the increased risk of colorectal cancer^8^, vegetarians also generally consume higher amounts of plant foods such as fruits, vegetables, and whole grains compared to meat-eaters^9, 10^, which might also contribute to lowering the risk of some site-specific cancers^8^. Evidence from two large cohorts which include a large proportion of vegetarians, the European Prospective Investigation into Cancer and Nutrition-Oxford (EPIC-Oxford) and the Adventist Health Study-2 (AHS-2), has suggested that vegetarians may have a lower risk of developing cancer (all types combined) in comparison to meat-eaters^11, 12^, but the evidence remains unclear for individual cancer sites^11, 13-16^. Moreover, the risk of cancer in those who do not consume meat but do eat fish (fish-eaters or pescatarians) may differ from that of meat-eaters; some evidence has suggested that fish-eaters may have a lower overall risk of cancer^11^, and a lower risk of colorectal cancer^11, 15^ than meat-eaters, but no differences have been reported for breast^13^ or prostate cancer risk^11, 14^. Despite the large number of vegetarians and fish-eaters in these cohorts (8,000 - 25,000 participants), power to detect an association for specific cancer sites may be limited due to relatively small numbers of cancer cases (∼5000 total cases) in these individual studies^11, 13-16^.

Any difference in cancer risk between diet groups may be due to differences in physiological characteristics, including adiposity. In western populations, vegetarians and fish-eaters have been shown to have lower body mass indices in comparison with the body mass index (BMI) of meat-eaters^17-19^ which is important for cancer risk because obesity is a known risk factor for several cancer sites^8^. Another hypothesized explanation for the lower risk of cancer observed among vegetarians and fish-eaters is the possible differences in hormone levels^20^, such as insulin-like growth factor-I (IGF-I) and testosterone, which may be related to their dietary intakes^20-22^. Hormone difference may be important as higher levels of IGF-I have been associated with higher risks of colorectal, breast, and prostate cancer^23^ and higher levels of free testosterone have been associated with prostate cancer^24^ and postmenopausal breast cancer.^25^

To further understand these relationships, we assessed the associations of diet groups with risks of all, colorectal, postmenopausal breast, and prostate cancer in the UK Biobank, which includes 10,000 fish-eaters, 8,000 vegetarians, and nearly 55,000 total incident cancer cases. We additionally aimed to assess the roles of BMI, circulating IGF-I, and calculated free testosterone as potential mediators of the observed associations between diet groups and cancer risk.

## Methods

### Study design and participants

Potential participants were first identified for the UK Biobank study using National Health Service (NHS) records, and 9.2 million eligible individuals, aged 40-70 and living within 25 miles of one of the assessment centres in the UK, were invited to participate in the study. Over 500,000 participants (5.5% response rate) consented to participate between 2006 to 2010^26^ and visited one of 22 assessments centres across England, Wales, and Scotland. A full description of the study protocol can be found on the UK Biobank website^26^.

The UK Biobank was approved by the NHS North West Multicentre Research Ethics Committee (16/NW/0274). All participants provided informed consent at recruitment, allowing for follow-up using data-linkage to health records.

### Exclusions

Participants were excluded from this analysis if they withdrew consent over the study period (n=871), had a prevalent cancer diagnosis at recruitment (excluding non-melanoma skin cancer International Statistical Classification of Disease (ICD-10) code: C44; n=29,504), their genetic sex was different from their reported sex (n=321), or they did not contribute any follow-up time (n=2; Supplementary Figure 1). Participants who responded as ‘do not know’ or ‘prefer not to say’ for all dietary questions regarding meat intake were also excluded from the analyses (n=282). This left a total of 472,337 participants, of whom 217,937 were males and 254,400 were females. For prostate cancer analyses women were excluded, and for postmenopausal breast cancer analyses women who were premenopausal at recruitment and did not reach the age of 55 over the follow-up time (n=16,222), and men, were excluded.

### Diet group classification

Diet groups were categorised using the touchscreen questionnaire completed at recruitment which asked participants about their frequency of consumption of processed meat, beef, lamb or mutton, pork, chicken, turkey or other poultry, and oily and non-oily fish. Participants chose a frequency of intake ranging from “Never” to “Once or more daily”. From these responses, participants were categorised into four diet groups (regular meat-eaters; low meat-eaters; fish-eaters; and vegetarians). Regular meat-eaters were participants who said they consumed processed, red meat (beef, pork, lamb), or poultry >5 times a week. Low meat-eaters were participants who reported consuming processed, red meat, or poultry ≤5 times a week. Fish-eaters were participants who reported that they never consumed red meat, processed meat, or poultry but ate oily and/or non-oily fish. Vegetarians were defined as participants who reported that they never consumed any meat or fish. The vegetarian group also included vegans who reported not consuming any meat, fish, dairy, or eggs (n=446).

### Covariates and biomarkers

The baseline touchscreen questionnaire also asked participants about sociodemographic, reproductive, and lifestyle factors. In addition, all participants had their blood drawn and anthropometric measurements, including height and weight, taken by a trained professional. Further information on covariate data collection and classification can be found in the **Supplementary Material**.

Non-fasting blood samples were provided by 99.7% of participants at recruitment and were shipped to the central processing laboratory at 4°C prior to serum preparation, aliquoting, and cryopreservation in the central working archive. Biochemistry markers were measured including insulin-like growth factor-I (IGF-I) and testosterone. Further description of the UK Biobank biomarker measurements can be found online^27^.

### Follow-up and outcome ascertainment

Data on cancer diagnosis were ascertained using a combination of records from the NHS Digital (cancer registry) and Public Health England for participants from England and Wales, NHS Central Register for participants from Scotland^28^ as well as the Hospital Episodes Statistics (HES) data for English participants and Scottish Morbidity Records (SMR) for Scottish participants (please see details in the Supplementary Methods). Using the World Health Organization’s ICD-10 codes, participants were classified as having an event if they had an incident diagnosis of cancer recorded as: all cancer (C00-97 excluding non-melanoma skin cancer: C44), colorectal cancer (C18-C20), breast cancer (C50), or prostate cancer (C61) or if no prior incident diagnosis was reported their primary underlying cause of death was the respective cancer. Participants contributed follow-up time from the date of recruitment until the date of the first cancer registration or cancer first recorded on death certificate, date of death, or last day of follow-up available from HES and SMR data (28^th^ February 2021 for England & Scotland). Cancer registry data were available until 31^st^ July 2019 for England and Wales, and 31^st^ October 2015 for Scotland, after this time only HES and SMR data were used for the follow-up of participants. For Welsh participants, hospital episode data did not extend past the cancer registry censoring date and therefore were not used. For breast cancer, analyses were restricted to postmenopausal breast cancer and women contributed follow-up time beginning when they turned 55 years of age, or their date at recruitment if they were categorised as being postmenopausal from questions asked at baseline (see **Supplementary Methods** for further details).

### Statistical analyses

Baseline characteristics of UK Biobank participants were summarised across diet groups for all participants, and separately for men and women.

Cox proportional hazards regressions were used, with age as the underlying time variable, to estimate hazard ratios (HR) and 95% confidence intervals (CI). Minimally adjusted models were stratified by sex (for all cancer and colorectal cancer analyses only) and age at recruitment (<45, 45-49, 50-54, 55-59, 60-64, ≥65 years) and adjusted for region at recruitment (North-West England, North-Eastern England, Yorkshire & the Humber, West Midlands, East Midlands, South-East England, South-West England, London, Wales, and Scotland).

Multivariable-adjusted Cox regression models for all analyses were further adjusted for height (eight sex-specific categories increasing by 5 cm, and unknown/missing (0.51%)), physical activity (low: 0-9.99, medium: 10-49.99, high: ≥ 50 metabolic equivalent of task-hours /week, and unknown/missing (4.04%)), Townsend deprivation index (quintiles from most deprived to least deprived, and unknown/missing (0.13%)), education (completion of national exam at 16, completion of national exam at 17-18, college or university degree, or other/unknown/missing (18.7%)), employment status (employed, retired, not in paid employment, or unknown (1.15%)), smoking status (never, former, light smoker: ≤15 cigarettes/day, medium smoker: 16-29 cigarettes/day, heavy smoker: ≥30 cigarettes/day, or missing/unknown (0.65%)), alcohol consumption (none drinkers, <1, 1-9.99, 10-19.99, ≥20 grams/day, or unknown/missing (0.73%)), ethnicity (White, Mixed race or other, Asian or British Asian, and Black or Black British, or missing/unknown (0.56%)), and diabetes status (no, yes, or unknown (0.53%)).

For colorectal cancer and for all cancer sites, multivariable models were further adjusted for female specific covariates: menopausal hormone therapy (MHT) use (no, former, current, or unknown (0.58%)), and menopausal status at recruitment (premenopausal, postmenopausal, or unknown (9.0%)). Moreover, for colorectal cancer, multivariable models were adjusted for non-steroidal anti-inflammatory drug use (NSAID; no reported use, irregular use, regular use of aspirin/ibuprofen). For prostate cancer, models were additionally adjusted for marital status (not living with a partner, living with a partner)^29^. For postmenopausal breast cancer, models were additionally adjusted for MHT use (same as above), age at menarche (≤12 years, 13 years old, ≥14 years, or unknown (22.5%)), parity and age at first birth (nulliparous, 1-2 children <25 years old, 3+ children <25 years old, 1-2 children 25-29.9 years old, 3+ children 25-29.9 years old, 1-2 children 30+ years old, 3+ children 30+ years old, or missing (0.3%)). In all models, the proportional hazards assumption was evaluated using Schoenfeld residuals, and no violations were observed.

We considered BMI as a potential confounder as well as a mediator. When BMI was considered as a potential confounder, BMI measured at recruitment was added to multivariable models (multivariable adjusted + BMI; <20, 20-22.49, 22.5-24.9, 25.0-27.49, 27.5-29.9, 30-32.49, 32.5-34.9, ≥35 kg/m^2^, or unknown/missing (0.57%)). Models assessing BMI as a mediator are explained below in the mediation analyses section.

To determine if there was heterogeneity in the associations of diet groups with cancer risk, and to assess the influence of confounder adjustments^30, 31^, *χ*^2^ statistics and p-values for including the diet group in the model were estimated using likelihood ratio tests (LRT) comparing a model without the diet groups variable to the model with the diet groups.

### Subgroup and sensitivity analyses

For all analyses, we assessed heterogeneity by subgroups of BMI (median: <27.5 and ≥27.5 kg/m^2^) and smoking status (ever and never) by using a LRT comparing the main model to a model including an interaction term between diet groups and the subgroup variable (BMI and smoking status). For colorectal cancer we further assessed heterogeneity by sex. For all cancer sites combined we additionally explored heterogeneity by smoking status, censoring participants at baseline who were diagnosed with lung cancer.

In sensitivity analyses, we excluded cases and participants who had less than 2 years of follow-up and all participants with missing data on covariates. We also examined associations separately in white participants because a large proportion of the vegetarians in this cohort are of South Asian ethnicity. Furthermore, we additionally adjusted for fruit and vegetable intake in the multivariable adjusted model (<3 servings/day, 3-3.99 servings/day, 4-5.99 servings/day, ≥6 servings/day, unknown) to control for this component of dietary intake as a proxy for a healthy diet. For prostate cancer analyses, we included in the multivariable adjusted model prostate specific antigen (PSA) testing (no PSA testing, had PSA test, or unknown) reported at baseline in all men and during follow-up from general practice records in a subsample (n=99,412 males; records available for participants until 31^st^ of May 2016 for England, 31^st^ of March 2017 for Scotland, and 31^st^ of August 2017 for Wales).

### Mediation analyses

If a significant association was observed between a diet group and a cancer outcome in the main analyses, we then further explored potential mediators that have been shown to be associated with diet groups^18, 20^ and were previously related to the cancer site of interest (BMI, IGF-I, and free testosterone)^24, 25^. To determine if differences in mediators were observed by diet group, we used multivariable linear regression to compare the selected biomarker measurements (IGF-I and free testosterone) and BMI across dietary groups, adjusting for potential confounders (see **Supplementary Methods**). We did not explore mediation if there was no significant difference in cancer risk between each diet groups and regular meat-eaters, or if the biomarker concentrations were not significantly different between diet groups. We explored mediation via BMI for all cancer, colorectal cancer, and postmenopausal breast cancer risk^8^, but not for prostate cancer due to its heterogeneous association with risk by stage and grade^32^ and as these data are not available in this cohort. For prostate cancer and postmenopausal breast cancer we also explored potential mediation via circulating concentrations of IGF-I and calculated free testosterone^24, 25^. We did not explore biomarker mediation for the all cancer - diet group associations as these biomarkers have not been associated with all cancer risk.

To assess for mediation, we used the inverse odds ratio weighting (IORW) method^33, 34^. This method aims to decompose associations between diet group mediated by the potential mediator (natural indirect effect [NIE]) and the estimated association of diet group with cancer risk not mediated by baseline BMI or biomarkers (natural direct effect [NDE]). The term “effect” is used here in concordance with the causal mediation literature but should not be interpreted as implying causality. To determine the proportion of the association between diet groups and cancer outcome mediated by the mediator of interest (e.g. BMI) we took the log of the indirect effect HR and divided it by the log of the total effect HR. Further details of the mediation analyses can be found in the **Supplementary Methods**.

All analyses were conducted using Stata version 17.0 (Stata Corp LP, College Station, TX). P-values were two-sided with p <0.05 being considered statistically significant.

## Results

Of the participants included in the analysis, 247,571 (52.4%) were classified as regular meat-eaters, 205,385 (43.5%) were low meat-eaters, 10,696 (2.3%) were fish-eaters and 8,685 (1.8%) were vegetarians. After an average of 11.4 years of follow-up, 54,961 incident cases of any type of cancer were diagnosed; 5,882 participants were diagnosed with colorectal cancer, 7,537 women were diagnosed with postmenopausal breast cancer, and 9,501 men were diagnosed with prostate cancer.

**Table 1** presents participants’ baseline characteristics across diet groups. Vegetarians and fish-eaters had a lower BMI, were younger, more likely to be never smokers, have a university/college degree, and report consuming less alcohol at recruitment compared to regular meat-eaters. Vegetarian men were also less likely to have had a PSA test in comparison to meat-eaters (**Table 1**). Supplementary Table 1 presents the baseline characteristics across diet groups stratified by sex. Both men and women fish-eaters and vegetarians had lower BMIs and were younger at recruitment in comparison to regular meat-eaters.

**Table 1.** Baseline characteristics across diet groups in UK Biobank

|  | Diet groups |  |  |  |
| --- | --- | --- | --- | --- |
|  | Regular meat-eaters | Low meat-eaters | Fish-eaters | Vegetarians |
| Number of participants | 247,571 | 205,385 | 10,696 | 8,685 |
| Age at recruitment- years, mean (SD) | 56.0 (8.2) | 56.9 (8.0) | 54.0 (8.0) | 53.0 (7.9) |
| Sex - Female | 114849 (46.4%) | 126165 (61.4%) | 7664 (71.7%) | 5722 (65.9%) |
| BMI - kg/m<sup>2</sup>, mean (SD) | 27.9 (4.9) | 27.0 (4.7) | 25.3 (4.3) | 25.7 (4.7) |
| Male height - cm, mean (SD) | 175.7 (6.8) | 175.4 (6.9) | 176.4 (6.9) | 175.5 (7.2) |
| Female height - cm, mean (SD) | 162.4 (6.3) | 162.5 (6.3) | 163.5 (6.4) | 162.1 (6.8) |
| Physical activity |  |  |  |  |
| Low | 72,811 (29.4%) | 59,752 (29.1%) | 2,430 (22.7%) | 2,371 (27.3%) |
| Moderate | 116,591 (47.1%) | 98,692 (48.1%) | 5,710 (53.4%) | 4,346 (50.0%) |
| High | 48,012 (19.4%) | 38,704 (18.8%) | 2,273 (21.3%) | 1,690 (19.5%) |
| Townsend deprivation index |  |  |  |  |
| Q1- Most affluent | 51,117 (20.6%) | 40,433 (19.7%) | 1,777 (16.6%) | 1,258 (14.5%) |
| Q5 - Most deprived | 48,227 (19.5%) | 41,627 (20.3%) | 2,385 (22.3%) | 2,216 (25.5%) |
| Ethnicity |  |  |  |  |
| White | 233,959 (94.5%) | 193,033 (94.0%) | 9,922 (92.8%) | 6,903 (79.5%) |
| Mixed other | 3,576 (1.4%) | 3,294 (1.6%) | 172 (1.6%) | 152 (1.8%) |
| Asian or British Asian | 4,114 (1.7%) | 5,054 (2.5%) | 369 (3.4%) | 1524 (17.5%) |
| Black or Black British | 4,218 (1.7%) | 3,295 (1.6%) | 167 (1.6%) | 48 (0.6%) |
| Education |  |  |  |  |
| National exam at 16 years | 41,764 (16.9%) | 34,271 (16.7%) | 1,180 (11.0%) | 1,099 (12.7%) |
| National exam at 17-18 years | 13,750 (5.6%) | 10,805 (5.3%) | 578 (5.4%) | 551 (6.3%) |
| Degree or college | 146,214 (59.1%) | 119,791 (58.3%) | 8,015 (74.9%) | 6,109 (70.3%) |
| Employment |  |  |  |  |
| In paid employment | 146,078 (59.0%) | 115,579 (56.3%) | 7,338 (68.6%) | 6,065 (69.8%) |
| Retired | 77,483 (31.3%) | 70,640 (34.4%) | 2,341 (21.9%) | 1,582 (18.2%) |
| Not in paid employment | 21,068 (8.5%) | 17,028 (8.3%) | 877 (8.2%) | 921 (10.6%) |
| Living with a partner - Yes | 187,545 (75.8%) | 141,711 (69.0%) | 6,930 (64.8%) | 5,771 (66.4%) |
| Smoking status |  |  |  |  |
| Never | 132,294 (53.4%) | 114,385 (55.7%) | 6,075 (56.8%) | 5,561 (64.0%) |
| Previous | 85,319 (34.5%) | 69,642 (33.9%) | 3,800 (35.5%) | 2,480 (28.6%) |
| Light smoker <15 cig/day | 7,594 (3.1%) | 6,299 (3.1%) | 290 (2.7%) | 210 (2.4%) |
| Medium smoker 15-29 cig/day | 10,101 (4.1%) | 6,644 (3.2%) | 157 (1.5%) | 139 (1.6%) |
| Heavy smoker 30+ cig/day | 10,418 (4.2%) | 7,432 (3.6%) | 333 (3.1%) | 252 (2.9%) |
| Alcohol intake g/day, mean (SD) | 19.9 (21.3) | 14.9 (16.6) | 13.6 (14.5) | 13.0 (16.1) |
| Diabetic - Yes | 15,603 (6.3%) | 10,748 (5.2%) | 290 (2.7%) | 465 (5.4%) |
| Prostate specific antigen test reported at baseline or in follow-up – Yes, Male only | 51,555 (38.8%) | 33,394 (42.2%) | 1,125 (37.1%) | 929 (31.4%) |
| **Female specific covariates** |  |  |  |  |
| Age at first menarche – years, mean (SD) | 12.6 (2.8) | 12.5 (2.9) | 12.5 (2.9) | 12.4 (3.3) |
| Menopause status |  |  |  |  |
| Premenopausal | 24,939 (21.7%) | 23,360 (18.5%) | 2,232 (29.1%) | 1,843 (32.2%) |
| Postmenopausal | 78,626 (68.5%) | 92,413 (73.2%) | 4,717 (61.5%) | 3,342 (58.4%) |
| Menopausal hormone therapy use |  |  |  |  |
| Never | 70,830 (61.7%) | 76,747 (60.8%) | 5,468 (71.3%) | 4,386 (76.7%) |
| Former | 34,149 (29.7%) | 38,960 (30.9%) | 1,590 (20.7%) | 9,66 (16.9%) |
| Current | 8,993 (7.8%) | 9,988 (7.9%) | 584 (7.6%) | 320 (5.6%) |
| Parity |  |  |  |  |
| Nulliparous | 17,671 (15.4%) | 25,569 (20.3%) | 2,330 (30.4%) | 1,736 (30.3%) |
| 1-2 children | 67,306 (58.6%) | 70,915 (56.2%) | 3,910 (51.0%) | 2,770 (48.4%) |
| 3+ children | 29,343 (25.5%) | 29,526 (23.4%) | 1,415 (18.5%) | 1,203 (21.0%) |
| Age at first birth – years, mean (SD) | 25.4 (4.6) | 25.2 (4.6) | 26.4 (5.1) | 26.0 (4.9) |

Percentages include missing values and therefore may not add up to 100%.
Values are N (%) unless otherwise indicated.
Abbreviations: BMI: body mass index; cig, cigarette; Q: quintile; SD: standard deviation.

The minimally adjusted models and sequential adjustments for the associations between diet groups and cancer risks are presented in Supplementary Table 2, and **Figure 1** shows the multivariable-adjusted models. In the multivariable-adjusted models (not including BMI), a vegetarian diet was associated with a lower risk of all cancer (HR: 0.86, 95% CI: 0.80-0.93), postmenopausal breast cancer (0.82, 0.68-0.99) and prostate cancer (0.69, 0.54-0.89; **Figure 1A**). Furthermore, compared to being a regular meat-eater, fish-eaters had a lower risk of all cancers (0.90, 0.84-0.96) and prostate cancer (0.80, 0.65-0.99), and low meat-eaters had a lower risk of colorectal cancer (0.91, 0.86-0.96; **Figure 1A**). When including BMI as a potential confounder, associations were slightly attenuated apart from prostate cancer which did not change (**Figure 1B**). For postmenopausal breast cancer, after adjustment for BMI the risk for vegetarians compared to regular meat-eaters was no longer statistically significant (0.87, 0.72-1.05; **Figure 1B**).

**Figure 1.**
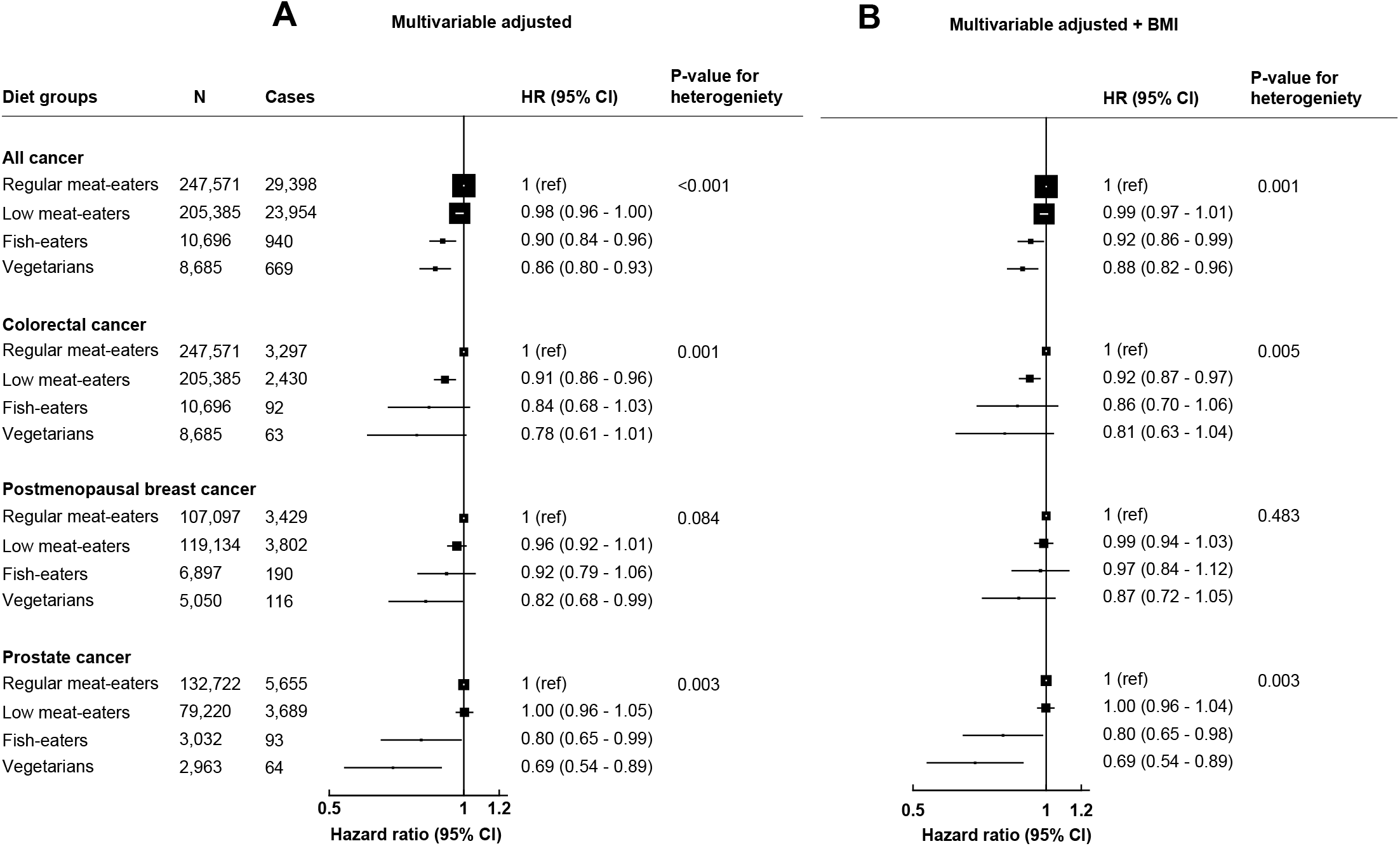
Multivariable adjusted hazard ratios (95% CI) for diet groups and risk of all cancer, prostate cancer, postmenopausal breast cancer and colorectal cancer not adjusting for BMI (A) and adjusting for BMI (B). Regular meat-eaters: red or processed meat or poultry >5 times a week. Low meat-eaters: red and processed meat or poultry ≤5 times per week. Fish-eaters: do not consume red, processed meat, or poultry but consumed fish. Vegetarians (including vegans): do not consume any meat or fish. All models used age as the underlying time variable and are stratified by sex (for only all cancer and colorectal cancer), age groups, and adjusted for region of recruitment, height, physical activity, Townsend deprivation index, education, employment status, smoking status, alcohol consumption, ethnicity, and diabetes status. Full details for each covariate are provided in the statistical analysis section in the main text. For all cancer and colorectal cancer analyses, models were further adjusted for menopausal hormone therapy use and menopausal status and colorectal cancer models are adjusted for non-steroid anti-inflammatory drug use. For prostate cancer models are further adjusted for marital status. For breast cancer model are further adjusted for menopausal hormone therapy use, age at menarche, and age at first birth/parity. Multivariable + BMI models further adjusts for BMI. *P*-value for heterogeneity from likelihood ratio tests for model fit comparing a model without diet groups, to a model including diet group. Abbreviations: BMI, body mass index; CI, confidence intervals; HR, hazard ratio; N, number of participants.

### Subgroup analyses

No evidence of heterogeneity was observed across BMI subgroups in the associations between diet groups and risk of all cancer, colorectal, postmenopausal breast, and prostate cancer (Supplementary Tables 3-6). For colorectal cancer, there was evidence of heterogeneity by sex (P_het_ =0.007), with male low meat-eaters, fish-eaters, and vegetarians having a lower risk of colorectal cancer (0.89, 0.83-0.95; 0.69, 0.47-1.01; 0.57, 0.36-0.91, respectively) in comparison to regular meat-eaters, whereas no significant association was observed across diet groups for females (Supplementary Table 4). For smoking status, some evidence of heterogeneity was observed in the association between diet groups and all cancer risk (p_het_=0.06); among ever smokers, the low meat-eaters, fish-eaters, and vegetarians had lower risks of all cancer sites than regular meat-eaters (0.97, 0.94-0.99; 0.86, 0.78-0.95; 0.79, 0.70-0.90, respectively), whereas these associations were non-significant for non-smokers (Supplementary Table 3). However, when censoring participants with lung cancer at baseline, the test for heterogeneity by smoking status became non-significant between diet groups (p_het_=0.22; Supplementary Table 3) although the association with diet group was only significant among smokers.

### Sensitivity analyses

Associations remained largely the same when analyses were restricted to participants of white European ancestry, and when participants with missing data were excluded (Supplementary Figure 2). When the participants who had an event or were censored in the first two years of follow-up were excluded, results remained mostly the same except that being a fish-eater was more strongly associated with a lower risk of prostate cancer (HR 0.69, 0.55-0.88) in comparison to regular meat-eaters (Supplementary Figure 3). In analyses additionally adjusted for intake of fruit and vegetables in the multivariable models, no changes in associations were observed (Supplementary Figure 3). For prostate cancer risk, when PSA testing was added to multivariable models the associations were not materially changed (Supplementary Table 2).

### Mediation analyses

Adjusted and relative means of BMI, IGF-I, and free testosterone across diet groups are shown in Supplementary Table 7. Explorations of potential mediators for significant diet-cancer associations are shown in **Table 2**. When we considered the potential of mediation via BMI in the associations of diet groups and risk of all cancer, this was not found to substantially mediate the observed associations (**Table 2**). For colorectal cancer risk, BMI was not found to mediate the observed lower risk in low meat-eaters compared with regular meat-eaters (**Table 2**); hormonal biomarkers were not explored due to no differences in concentrations observed between regular and low meat-eaters (Supplementary Table 7). For postmenopausal breast cancer risk, BMI was found to be a potential mediator for the observed difference in the risk between vegetarians and regular meat-eaters, with a decomposed HR^NIE^ of 0.83 (95% CI: 0.63-1.08) explaining nearly 93% of the lower risk observed in vegetarian women (**Table 2**). When IGF-I was explored independently and after adjusting for BMI, a HR^NIE^ of 0.91 (95% CI: 0.73-1.15) was observed. For prostate cancer risk, IGF-I and free testosterone concentrations did not seem to mediate the observed difference in risk between vegetarians and regular meat-eaters and free testosterone was not found to mediate the difference in risk between fish-eaters and regular meat-eaters (**Table 2**).

**Table 2.** Summary of estimated direct effect, indirect effect, and total effect using potential mediators for the association of diet groups in comparison to regular meat-eaters and risk of all cancer, colorectal cancer, postmenopausal breast cancer, and prostate cancer risk

| Potential mediators (Hazard ratio; 95% CI) |  |  |  |
| --- | --- | --- | --- |
| **All cancer** | **Mediation through BMI** | **Mediation through IGF-I <sup>a</sup>** | **Mediation through free testosterone <sup>a</sup>** |
| Low meat-eaters versus regular meat-eaters | (n=450,412) |  |  |
| Total effect | 0.99 (0.96 – 1.00) |  |  |
| Natural indirect effect | 0.99 (0.98 – 1.00) |  |  |
| Natural direct effect | 0.99 (0.95 – 1.00) |  |  |
|  | Mediation through BMI |  |  |
|  | (n=256,727) |  |  |
| Fish-eaters versus regular meat-eaters |  |  |  |
| Total effect | 0.90 (0.83 – 0.97) |  |  |
| Natural indirect effect | 0.99 (0.90 – 1.01) |  |  |
| Natural direct effect | 0.90 (0.83 – 0.98) |  |  |
|  | Mediation through BMI |  |  |
|  | (n=254,709) |  |  |
| Vegetarians versus regular meat-eaters |  |  |  |
| Total effect | 0.86 (0.78 – 0.96) |  |  |
| Natural indirect effect | 0.94 (0.81 – 1.08) |  |  |
| Natural direct effect | 0.92 (0.77 – 1.09) |  |  |
| **Colorectal cancer** | Mediation through BMI |  |  |
|  | (n=450,412) |  |  |
| Low meat-eaters versus regular meat-eaters |  |  |  |
| Total effect | 0.91 (0.85 – 0.97) |  |  |
| Natural indirect effect | 1.00 (0.98 – 1.02) |  |  |
| Natural direct effect | 0.91 (0.86 – 0.97) |  |  |
| **Postmenopausal breast cancer** | Mediation through BMI | Mediation through IGF-I | Mediation through free testosterone |
|  | (n=111,574) | (n=103,853) | (n=93,662) |
| Vegetarians versus regular meat-eaters |  |  |  |
| Total effect | 0.82 (0.68 – 0.99) | 0.86 (0.71 – 1.05) | 0.86 (0.71 – 1.05) |
| Natural indirect effect | 0.83 (0.63 – 1.08) | 0.91 (0.73 – 1.15) | 1.06 (0.76 – 1.38) |
| Natural direct effect | 0.99 (0.79 – 1.23) | 0.94 (0.70 – 1.21) | 0.81 (0.62 – 1.05) |

| **Prostate cancer<sup>b</sup>** |  | Mediation through IGF-I<br>(n=126,538) | Mediation through free testosterone<br>(n=116,087) |
| --- | --- | --- | --- |
| Vegetarians versus regular meat-eaters |  |  |  |
| Total effect |  | 0.71 (0.56 - 0.92) | 0.71 (0.56 - 0.92) |
| Natural indirect effect |  | 1.10 (0.77 - 1.56) | 0.99 (0.67 - 1.48) |
| Natural direct effect |  | 0.64 (0.50 - 1.01) | 0.71 (0.51 - 1.01) |
| Fish-eaters versus regular meat-eaters |  |  | Mediation through free testosterone<br>(n=116,186) |
| Total effect |  |  | 0.80 (0.65-0.99) |
| Natural indirect effect |  |  | 0.95 (0.70-1.29) |
| Natural direct effect |  |  | 0.86 (0.56-1.32) |

All models used age as the underlying time variable and are stratified by sex (for only all cancer and colorectal cancer) and age groups at recruitment, and adjusted for region of recruitment, height, physical activity, Townsend deprivation index, education, employment status, smoking status, alcohol consumption, ethnicity, diabetes status, and body mass index (except when it was considered a potential mediator). For all cancer and colorectal cancer, models are further adjusted for menopausal hormone therapy use and menopausal status. Colorectal cancer models are adjusted for non-steroid anti-inflammatory drug use. For prostate cancer models are further adjusted for marital status. For breast cancer model are further adjusted for menopausal hormone therapy use, age at menarche, and age at first birth/ parity. Full details for each covariate are provided in the statistical analysis section in the main text.
Natural indirect effect represents the estimated association of diet group and cancer outcome through the potential mediator.
Natural direct effect represents the estimated association of diet group and cancer outcome not through the potential mediator.
Models exclude participants with missing values for mediator(s).
<sup>a</sup> Models are adjusted for BMI.
<sup>b</sup> BMI not assessed as a mediator with total prostate cancer risk. Association of IGF-I and free testosterone presented as both hormones have been associated with prostate cancer risk. IGF-I concentrations not assessed for fish-eaters as no difference in concentrations in comparison to regular meat-eaters was observed.
Abbreviations: BMI: body mass index; CI, confidence intervals; IGF-I, insulin like growth factor-I.

## Discussion

In this large British cohort, being a low meat-eater, fish-eater, or vegetarian was associated with a lower risk of all cancer sites when compared to regular meat-eaters. We also found a lower risk of colorectal cancer amongst low meat-eaters, a lower risk of postmenopausal breast cancer risk in vegetarian women, and a lower risk of prostate cancer among vegetarian men. The lower risk of postmenopausal breast cancer in vegetarians may be largely a result of vegetarians having a lower BMI than regular meat-eaters, with possibly some further impact due to vegetarian women in this population having slightly lower circulating IGF-I concentrations.

### All cancer

In this study, vegetarians, fish-eaters, and low meat-eaters all had a lower risk of developing all cancer in comparison to regular meat-eaters. It is important to consider that although some cancers may have similar aetiologies, some cancer sites may not be associated with dietary or nutritional factors and that using all cancer incidence as an outcome may crudely capture other lifestyle factors, outside of diet, that may be associated with cancer risk; therefore, these results should be interpreted carefully. In the two largest previous prospective studies following vegetarians, both EPIC-Oxford and AHS-2 found that being a vegetarian was associated with a 10% and 8% lower risk of all cancer than being a meat-eater, respectively, after adjusting for lifestyle risk factors and BMI^11, 12^. Fish-eaters in EPIC-Oxford had a lower risk of developing all cancer^11^ but no association with risk for all cancer was observed for fish-eaters in comparison to meat-eaters in AHS-2^12^. In the current analysis, we observed some evidence of heterogeneity by smoking status, and when we removed lung cancer from all cancer cases, significant associations were only observed across diet groups for the ever smoker subgroup. Therefore, the differences observed between diet groups for all cancer outcomes combined may be due to residual confounding by differences in other lifestyle factors, such as smoking.

### Colorectal cancer

The risk of colorectal cancer was lower in low meat-eaters in comparison to regular meat-eaters whereas there was no significant difference for fish-eaters and vegetarians, potentially due to lack of power because the point estimates suggested lower risks in both these non-meat-eating diet groups. In both EPIC-Oxford and AHS-2, being a fish-eater was associated with a lower risk of colorectal cancer in comparison with meat-eaters, whereas no association was observed for being vegetarian and risk of colorectal cancer compared to regular meat-eaters^11, 15^. We also observed heterogeneity by sex, in that significant inverse associations were observed with risk across diet groups in men, when compared to regular meat-eaters, but not for women. This may in part be due to dietary differences between sexes, however the number of colorectal cancer cases in some diet groups was too small to draw a clear conclusion. The intake of processed meat has been evaluated to be a definite cause of colorectal cancer^35^ and red meat as a probable cause of colorectal cancer^36, 37^. This is likely to at least in part explain the lower risk of colorectal cancer in low meat-eaters and mechanisms suggested including chemicals in meat such as nitrosamines^37^. Overweight and obesity increase the risk for colorectal cancer^38, 39^ but in mediation analyses BMI did not appear to mediate the association observed between low meat-eaters and regular meat-eaters.

### Postmenopausal breast cancer

A borderline significantly lower risk for postmenopausal breast cancer was observed for vegetarian women, which appeared to be largely due to their lower BMI as evidenced in mediation analyses and the attenuation of estimates when analyses were adjusted for BMI. We also observed a small potential effect for mediation for lower risk of postmenopausal breast cancer for vegetarians through lower IGF-I concentrations, perhaps influenced by the inclusion of vegans in this group^22^. To date, studies have reported a non-significantly lower risk of breast cancer for women following a vegetarian or pescatarian diet with or without adjustment for BMI^11, 13, 16, 40^, which may be due to lack of power to detect modest associations in individual studies. Breast cancer is a heterogeneous disease, with differing risk factors by menopausal status and hormone receptor status^41^. BMI is robustly associated with higher postmenopausal breast cancer risk, probably due to higher circulating oestrogen derived from adipose tissue^41^. As such, being vegetarian would be expected to confer a lower risk of postmenopausal breast cancer in comparison to meat-eaters because vegetarians generally have a lower BMI, but whether BMI is a confounder or a mediator for this association is not clear; if vegetarians have a lower BMI because of their diet then BMI would be a mediator, but if vegetarians have a lower BMI that is not due to their dietary intake but rather due to other non-dietary lifestyle factors (e.g. physical activity) then BMI would be considered to be a confounder.

Previous research has also suggested that vegetarian women are less likely to attend breast cancer screening and use MHT^42^, and thus the association may also be due to detection bias and residual confounding from MHT or other cofounders. As data on breast cancer screening during follow-up were not available in this cohort, adequate adjustment for screening attendance could not be made.

### Prostate cancer

The risk of prostate cancer was lower in men who were vegetarians or fish-eaters in comparison to regular meat-eaters, but no differences in risk were observed for low meat-eaters. Previous analyses in the EPIC-Oxford cohort found a non-significantly lower risk of prostate cancer for British vegetarians and fish-eaters in comparison to meat-eaters^11^. In the AHS-2 study, no difference was found for vegetarians or fish-eaters, whereas being vegan was associated with a 35% lower risk of prostate cancer (based on 1079 cases in the cohort of which only 59 were in vegans)^14^. To date, no established dietary risk factor has been found in relation to prostate cancer risk, however, there is some evidence which suggests that higher intake of dairy products, and possibly milk specifically, may increase the risk of prostate cancer^43^. This association has been proposed to be mediated through IGF-I^21, 44^, a hormone shown to be positively associated with both milk intake and prostate cancer risk^24, 45^. In this cohort, slightly lower IGF-I concentrations have been observed in vegetarians compared to regular meat-eaters^20^, and IGF-I has also been associated with prostate cancer risk^24^; however, the difference in IGF-I concentrations between these diet groups is small and may not confer a substantial difference in prostate cancer risk. As might be expected, in mediation analyses, the estimates were imprecise and there was no evidence that the difference in IGF-I concentrations between diet groups mediates the observed associations with cancer risk.

In this cohort vegetarian men were less likely to have had a PSA screening test than meat-eaters at recruitment, therefore they might have a higher risk of undetected or later detection of prostate cancer which translates into a lower risk of being diagnosed with prostate cancer during the follow-up period. Similarly, other cohorts have reported that vegetarian men were less likely to have had a PSA test^42, 46^. When data at recruitment and available general practice records during follow-up were assessed for PSA testing in UK Biobank, 40% of regular meat-eaters and 37% of vegetarians reported having had a PSA test (although general practice records were only available for half of the participants) after adjusting for age differences. Adding PSA screening in the multivariable-adjusted model did not attenuate the estimates, suggesting the differences in PSA screening in vegetarians or fish-eaters in comparison to regular meat-eaters does not explain the observed associations, but other differences in attendance for medical examinations could possibly also contribute. Due to unavailable data in UK Biobank, we were also unable to assess associations by tumour subtypes which may be aetiologically different^32^. Considering the substantial difference in risk we observed for vegetarian men, differences in detection and residual confounding, as well as chance, may contribute to this observed difference.

### Interpretation of results: role of confounding and mediation

The role of residual and unmeasured confounding must be considered when interpreting the findings from this study. Vegetarians and fish-eaters differ from meat-eaters in many non-dietary lifestyle factors such as lower smoking and alcohol consumption, and higher physical activity^47^. Although relevant potential confounders were added to the multivariable models to adjust for these differences, imperfect measurements and/or changes in these confounders over time may result in incomplete adjustment for these variables. For example, the evidence of heterogeneity by smoking status when looking at all cancer as an outcome suggested that residual confounding by smoking may operate.

Differences in BMI between diet groups have also been suggested to explain the lower cancer incidence observed amongst vegetarians^17^, however, when BMI was considered as a potential confounder and mediator, the difference between BMI by diet groups only slightly attenuated the estimates, with the exception of postmenopausal breast cancer. Whether differences in BMI by diet group is due solely to their diet or other lifestyle factors remains unclear, making it difficult to tease out whether BMI mediates or confounds the associations between diet group and cancer risk.

### Strengths and limitations

Strengths of this study include the prospective nature and moderately long follow-up time of participants. Data-linkage to health records was used to determine cancer diagnoses which minimises misclassification and loss to follow-up of participants. The UK Biobank study also gathered data on an array of potential confounders and biochemical biomarkers among participants, thus we were able to adjust the models for potential confounding as well as conduct mediation analysis exploring potential mediators between diet groups and cancer risk. When analyses excluded the first two years of follow-up, the results remained largely the same, reducing the chance that these associations are due to reverse causality.

There are some limitations to consider in these analyses. Although there were many cancer cases accrued during the follow-up period, these analyses may still be underpowered to detect moderate associations due to the relatively low numbers of cancer cases among vegetarians and fish-eaters in this cohort. We also used hospital admissions data to follow-up participants after 2015 in Scotland and 2019 in England because cancer registry data were not available in UK Biobank after this date, which may result in some missing cancer cases and relatively later dates of diagnosis. As detailed above, the results may be influenced by unmeasured and residual confounding, as well as chance with numerous comparisons, and causality cannot be confirmed. Misclassification of diet may also be possible, as participants may have underreported their intake or changed their diet over the follow-up period, possibly resulting in attenuation of the risk estimates. The mediation analyses only explored three potential mediators, and other possible mediating factors such as other biomarkers relevant for cancer, such as oestradiol, were not available. Moreover, baseline BMI and biomarkers were used to assess mediation, and therefore these measures may not represent BMI during the follow-up and long-term biomarker concentrations, although correlations with repeat measures of BMI and biomarkers showed high agreement^48^. As well, we used the IORW method to explore mediation with bootstrapping CIs, which may make estimates less statistically efficient in comparison to parametric methods, but the IORW has the advantage that it can be applied in survival analysis and provides estimates of the amount mediated for the mediators of interest. The UK Biobank has been shown to have a healthier risk profile than the UK population^49^ and only included British participants most of whom are of white European ancestry (94%); this may limit generalizability. However, the risks estimated may still be valid to estimate relative differences for risk-factor disease associations^50^.

In conclusion, this study found that being a low meat-eater, fish-eater, or vegetarian was associated with a lower risk of all cancer, which may be a result of dietary factors and/or non-dietary differences in lifestyle such as smoking. Low meat-eaters had a lower risk of colorectal cancer, vegetarian women had a lower risk of postmenopausal breast cancer, and men who were vegetarians or fish-eaters had a lower risk of prostate cancer. BMI was found to potentially mediate or confound the association between vegetarian diets and postmenopausal breast cancer. It is not clear if the other associations are causal or a result of differences in detection between diet groups or unmeasured and residual confounding. Future research assessing cancer risk in cohorts with large number of vegetarians is needed to provide more precise estimates of the associations and to explore other possible mechanisms or explanations for the observed differences.

## Data Availability

UK Biobank is an open access resource. Bona fide researchers can apply to use the UK Biobank dataset by registering and applying at http://ukbiobank.ac.uk/register-apply/.

## Acknowledgements

This work has been conducted using the UK Biobank Resource under Application Number 24494 and we wish to express our gratitude to the participants and those involved in building the resource. UK Biobank is an open access resource. Bona fide researchers can apply to use the UK Biobank dataset by registering and applying at http://ukbiobank.ac.uk/register-apply/.

