## Supplementary Material for "Risk of cancer in regular and low meat-eaters, fish-eaters, and vegetarians: a prospective analysis of UK Biobank participants"

### Supplementary Methods

#### Covariate classification

*Region*

Region of participants were grouped based on the recruitment centre they attended. A total of 10 regions were used corresponding approximately to the areas covered by the assessment centre: London (assessment centres: St Bartholomew's Hospital, Hounslow, Croydon), Wales (assessment centres: Swansea, Wrexham, Cardiff), North-West England (assessment centres: Stockport, Manchester, Liverpool, Bury), North-East England (assessment centres: Newcastle, Middlesbrough), Yorkshire (assessment centres: Leeds, Sheffield), West Midlands (assessment centres: Stoke, Birmingham), East Midlands (assessment centre: Nottingham), South-East England (assessment centres: Oxford, Reading), South-West England (assessment centre: Bristol), Scotland (assessment centres: Glasgow, Edinburgh).

*Height*

Participants were grouped into eight sex-specific categories for height. For females, categories were: <150, 150-154.9, 155-159.9, 160-164.9, 165-169.9, 170-174.9, 175-179.9, ≥180 cm. For males, categories were: <160, 160-164.9, 165-169.9, 170-174.9, 175-179.9, 180-184.9, 185-189.9, ≥190 cm. From this, males and females were combined into one height variable with categories going from 1 to 8, and a missing category for participants with missing data.

*Body mass index (BMI)*

Both height and weight were measured at the baseline visit and were used to determine participant's BMI. BMI was calculated by taking the participants measured weight in kilograms and dividing it by the participants squared standing height in metres. Individuals with missing data were coded into a missing category. Participants were categorised as follows: <20, 20-22.49, 22.5-24.99, 25.0-27.49, 27.5-29.99, 30-32.49, 32.5-34.99, ≥35 $kg/m^2$ and unknown/missing category.

*Alcohol intake*

Participants were asked on the baseline questionnaire at recruitment how often they drank alcohol with the possible responses being: “daily or almost daily”, “three or four times a week”, “once or twice a week”, “one to three times a month”, “special occasions only”, “never”, or “prefer not to answer”. Participants were also asked about their weekly and monthly intake of pints of beer, glasses of red wine, glasses of white wine/champagne, glasses of fortified wine, measures of spirits/liqueurs and glasses of other alcohol. A pint of beer was assumed to contain 20 grams of alcohol, and all other drinks contained 10 grams of alcohol. We then summed their total weekly and monthly consumption of alcohol accordingly. If the participant reported “do not know” or “prefer not to answer” to one of these questions on weekly or monthly consumption, they were coded as missing, except for “other alcohol”, in which case we assigned them 0 grams from other alcohol. We used participants reported weekly consumption of alcohol, if this was unknown (due to the participant reporting “do not know” or “prefer not to answer” for one or more of the relevant questions, except for “other alcohol”) we used monthly consumption, if available. To get an estimated daily total, we divided weekly consumption by 7 (or monthly consumption by 30.4375). Alcohol consumption was categorised as <1 g/day, 1-9.99 g/day, 10-19.99 g/day, and ≥20 g/day, never or unknown. For participants who had unknown grams/day of alcohol but who reported consuming alcohol intake on “special occasions”, we assigned them to the category of “<1 g/day”.

*Physical activity*

Physical activity was determined from questions on the touchscreen questionnaire which asked about walking, moderate physical activity, and vigorous physical activity. These were used to estimate excess metabolic equivalent (MET)-hours/week of physical activity during work and leisure time. For each of the three activity categories (walking, moderate physical activity and vigorous physical activity), participants were asked how many days in a typical week they did each of the activities for 10 minutes or more. For each category, participants who entered one or more days were then asked how many minutes they spent doing those activities on a typical day. For each activity category, the number of reported days was multiplied by the number of reported minutes on a typical day to generate duration of activity in minutes per week. Activity on a typical day of 1260 min per week (equivalent to an average of 3 hours per day) were truncated at 1260. Total MET values for each category from the International Physical Activity Questionnaire short form were: 3.3 for walking, 4.0 for moderate physical activity and 8.0 for vigorous physical activity. Excess MET values were therefore 2.3 for walking, 3.0 for moderate physical activity and 7.0 for vigorous physical activity. Excess MET-hours per week were calculated by multiplying the excess MET value for each activity by the duration of activity in hours per week.

*Townsend deprivation index*

Townsend deprivation index was based on the preceding national census output areas. Each participant was assigned a score in correspondence to the output area in which their postcode was located. From this, participants were categorised into quintiles from most deprived to least deprived and a missing category where postcode information was not provided.

*Smoking status*

Smoking was determined from questions from the recruitment questionnaire. Participants were asked “Do you smoke tobacco now?” and “in the past, how often have you smoked tobacco?” to determine their smoking status. Smokers were further divided based on how many cigarettes they said they smoked on average per day from the question “About how many cigarettes do you smoke on average each day?”. If participants did not respond to how many cigarettes they had on average each day, they were categorised into the missing group as they could not be accurately categorised.

*Ethnicity*

Ethnicity of participants was determined from questions in the touchscreen questionnaire “What is your ethnic group?”. Options included: White, Mixed race or other, Asian or Asian British, Black or Black British, Chinese, and other ethnic group. From this, participants were grouped into five categories: White, Mixed race or other, Asian or British Asian, and Black or Black British, or missing/unknown.

*Education*

For education, participants were asked “Which of the following qualifications do you have?” being able to select more than one. Possible answers were: College or University degree; A levels/AS levels or equivalent (General Certificate of Education (GCE) Advanced Level); O levels/ General Certificate of Secondary Education (GCSEs) or equivalent; Certificate of Secondary Education (CSEs) or equivalent; National vocational qualification (NVQ) or Higher National Diploma (HND) or Higher National Certificate (HNC) or equivalent; Other professional qualifications example: nursing, teaching; None of the above; Prefer not to answer. We grouped participants into the following categories, based on their highest reported level of education: (College or University degree, vocational qualifications (other professional qualifications/NVQ or HND or HNC), optional national exams at ages 17 to 18 years (A levels/AS levels), national exams at age 16 years (O levels/GCSEs/CSEs), none of the above, unknown (prefer not to answer)).

*Employment status*

Employment status at recruitment was assessed by asking participants “Which of the following describes your current situation?” in which they selected answers which were applicable to them including “In paid employment or self-employed”, “retired”, “looking after home and/or family”, “unable to work because of sickness or disability”, “unemployed”, “doing unpaid or voluntary work”, “full or part time student”, “none of the above” or “prefer not to answer”. Participants were defined as being “in paid employment” if they responded they were in paid employment or self-employed, “retired” if they responded they were retired, and “not in paid employment” if they reported being unemployed, inability to work, being a student, or having caring responsibilities for their family. Finally, an unknown/missing category consisted of participants who did not respond, said they prefer not to answer, or answered none of the above to the options in the question.

*Diabetes status*

Participant’s diabetes status was determined using multiple questions from recruitment. First, from the question “Has a doctor ever told you that you have diabetes?” participants were classified as “yes”, “no” or “unknown” based on their response. As well, participants who reported using metformin or insulin at recruitment were considered having diabetes and included in the "yes - diabetes" category. Finally, if a participant had a measured glycated hemoglobin (HbA1c) of ≥ 48 mmol/mol at recruitment, they were defined as having diabetes and included in the "yes - diabetes" category.

*Marital status*

Living with a partner was derived from a question asked at recruitment in which participants answered if they lived with anyone else in their household. If the participants reported to be living with a husband, wife, or partner in their household they were classified as "living with a partner" if the participant reported to be living with any other person or alone, they were categorised as "not living with a partner".

*Non-steroid anti-inflammatory drugs*

Non-steroid anti-inflammatory drug (NSAID) use was determined based on the medications reported at recruitment. Participants were categorised into three groups "Non-users", "irregular NSAID users", and "regular users of aspirin or ibuprofen". Participants were categorised as "regular users of aspirin or ibuprofen" if they responded to taking ibuprofen or aspirin regularly. Participants were categorised into irregular users if they responded to taking any classification of NSAID at recruitment. If no use of NSAIDs were reported at recruitment, participants were categorised as non-users.

*Vegetable and fruit intake*

For total vegetable intake, participants were asked to enter the number of heaped tablespoons of cooked vegetables and salad/raw vegetables eaten per day or select "less than one", "do not know" or "prefer not to answer". Two heaped tablespoons of vegetables were counted as a serving. For fresh fruit, participants were asked to enter the number of pieces of fresh fruit and dried fruit (with examples given as to what constitutes a piece eaten per day) or select "less than one", "do not know" or "prefer not to answer". One piece of fresh fruit (one apple, one banana, 10 grapes etc.) was counted as a serving. From this, servings of both vegetables and fruit were combined and participants were categorised as consuming <3 servings/day, 3.00-3.99 servings/day, 4.00-5.99 servings/day, ≥6.00 servings/day.

*Prostate specific antigen testing*

Prostate specific antigen (PSA) testing was determined from baseline reports from male participants as well as follow-up through general practice records. At recruitment men were asked if they ever had a PSA test in the past. From this, they answered “yes”, “no” and “do not know”. Additionally, 99,412 men had information from general practice records during follow-up (records available for participants until 31st of May 2016 for England, 31st of March 2017 for Scotland, and 31st of August 2017 for Wales). From the general practice records, men with a recorded PSA test in the general practice record were coded as having a PSA test. From both reported at baseline and the subsample of men with follow-up general practice records, men were coded as “no record of having a PSA” test, “had a PSA test”, or “unknown” if men reported “do not know” at recruitment and no general practice records were available.

<u>Women specific covariates</u>

All men were put into a separate category for all women-specific covariates when analyses included both men and women.

*Menopausal hormone therapy*

For women, use of menopausal hormone therapy (MHT) was categorised as “current user”, “former user” and “never user” or “unknown/missing” based on the questions asked about MHT use in the touchscreen questionnaire at recruitment. Women were asked “Have you ever used hormone replacement therapy?” and if they answered yes: “How old were you when you last used HRT?”. Women were asked to enter their age when they last used HRT or could choose “Still taking HRT”, or they could select “prefer not to answer” or “do not know”.

*Menopausal status at recruitment*

Menopausal status was first determined by multiple questions asked in the baseline questionnaire. Women were defined as being pre-menopausal if they:

- Answered “no” to the question regarding having gone through menopause, or
- Reported they were “not sure” or did not respond to if they had gone through menopause and:

    - Were <50 years of age, did not have a bilateral oophorectomy/hysterectomy, and reported they were not using menopausal hormone therapy.
    - Were <50 years of age, reported they were menstruating today, and did not have a bilateral oophorectomy/hysterectomy.

Women were defined as post-menopausal if they:

- Answered "yes" to having gone through menopause
- Answered "not sure" or did not answer if they had gone through menopause and:
    - were ≥55 years of age, or
    - had a bilateral oophorectomy

Women were defined as their menopausal status being unknown if:

- Answered "no" to having gone through menopause and:
    - Did not answer no to using HRT, or
    - Did not answer no to having a bilateral oophorectomy, or
    - Did not answer no to having a hysterectomy, or
    - Were 50-54.9 years of age.

Menopause status defined at recruitment was used as the covariate in models for all cancer and colorectal cancer. In our main analyses of postmenopausal breast cancer, women defined as postmenopausal at recruitment were included in this analysis. Moreover, we included women who were categorised as unknown or premenopausal at the age they turned 55 years and were entered into the analysis at this time and followed until the end or censor date or date of event. The age of 55 was used as ~98% of females would have undergone menopause by this age[1].

*Parity and age at first birth*

Parity was defined by the recruitment question of "how many children have you given birth to?". Women who said they had given birth were asked how old they were in years when they gave birth to their first child. Based on these responses, women were categorised into groups of 0 children, 1-2 children <25 years of age, or 3+ children <25 years of age, 1-2 children 25-29.9 years of age, or 3+ children 25-29.9 years of age, 1-2 children 30+years of age, or 3+ children

30+ years of age or unknown if the participants responded, “do not know” or “prefer not to answer”.

*Age at menarche*

Women were categorised based on the recruitment question “How old were you when your period started”. From this, women were categorised into age groups as: ≤12 years, 13 years old, ≥14 years old. If a participant responded “prefer not to answer” or “do not know” they were categorised into an unknown group.

### Outcome ascertainment

Participants were first followed using National Health Service (NHS) Digital and Public Health England for participants from England and Wales and NHS Central Register for participants from Scotland. Follow-up was available until July 31st, 2019 for England and Wales and October 31st, 2015 for Scotland from these databases. After these dates, participants from England and Scotland were followed using Hospital Episode Statistics admissions and Scottish Morbidity Records which covered time periods until 28th February 2021 for England and Scotland. Participants information on primary and secondary cause for in-patient hospital admissions were used to determine an incident cancer diagnosis. If an ICD-10 code indicated their primary or secondary reason for visit was a cancer diagnosis, they were indicated as being a case at the earliest date indictaed. First date of cancer incidence (excluding non-melanoma skin cancer: ICD-10: C44) from either data linkage source was used as the diagnosis date and incident cancer site.

### Mediation analysis

To determine potential mediators, we explored the association of BMI, IGF-I, and free testosterone across diet groups to assess if there was significant heterogeneity by diet groups. We used multivariable linear regression models adjusted for age, sex, region of recruitment, height, physical activity, Townsend deprivation index, education, employment status, smoking status, alcohol consumption, ethnicity, diabetes status, menopausal hormone therapy use, menopausal status, and body mass index (except when BMI was the outcome of interest) to assess biomarker and BMI associations across diet groups.

Mediation analyses were conducted if an association was observed between a diet group and all cancer risk or cancer specific site in the main analyses. Specifically, we examined potential mediation for all cancer risk for vegetarians versus regular meat-eaters, fish-eaters versus regular meat-eaters and low meat-eaters versus regular meat-eaters via BMI[2, 3]. For colorectal cancer risk, we examined potential mediation via BMI for low meat-eaters versus regular meat-eaters. Biomarkers were not assessed for mediation for colorectal cancer as no differences were observed between regular and low meat-eaters (see Supplementary Table 7). For postmenopausal breast cancer risk, we explored mediation for vegetarians versus regular meat-eaters via BMI, IGF-I, and free testosterone (estimated using a formula based on the law of mass action from measured total testosterone, sex-hormone binding globulin, and albumin concentrations[4]), for postmenopausal women. If a woman was missing a value for testosterone due to a very low serum concentration reported, the concentration for these participants was set to three-quarters of the minimum reportable value (0.35 nmol/L; n=23,494) and free testosterone was calculated from this. For prostate cancer risk we examined potential mediation for vegetarians versus regular meat-eaters via IGF-I and free testosterone, as these hormones have been associated with higher prostate cancer risk[5]. We also explored mediation via free testosterone for the fish-eater versus regular meat-eater analysis but not IGF-I as no differences in IGF-I concentrations was observed between fish-eaters and regular meat-eaters. BMI was not explored as a potential mediator for prostate cancer as BMI is heterogeneously associated with prostate cancer risk[6]. Specifically, BMI may be a risk factor for prostate cancer death and aggressive disease but has been inversely associated with total prostate cancer and non-aggressive disease risk[6, 7].

To assess for potential mediation, we used the inverse odds ratio weighting (IORW) method[8, 9] to estimate mediation of diet group through BMI or biomarkers. The IORW method was used as it is generally more flexible in estimating mediation as it does not require specification of regression models for each mediator of interest (like other traditional methods used for mediation[10, 11]), as well, it does not assume there is no exposure-mediator interactions, and can easily be applied to survival analyses, specifically Cox regression. The IORW evades these issues by condensing the relationship between exposure and mediators of interest into a single odds ratio (OR), thus an OR for the exposure conditional on all mediators is equivalent to the OR for the mediators, conditional on exposure. This OR can then be applied as a weight to a regression of the outcome on the exposure, thus estimating the natural direct effect (NDE) estimate as the pathway through which the mediators is deactivated. As with all mediation analyses, key assumptions are made using the IORW method, namely 1) there is no unmeasured exposure – mediator confounding, 2) there is no unmeasured mediator - outcome confounding, 3) there is no unmeasured exposure – outcome confounding, 4) there are no mediator-outcome confounders that is itself affected by the exposure[10].

The weight of mediators was estimated from logistic regression model where the binary diet group (i.e. vegetarians – regular meat-eaters; fish-eaters – regular meat-eaters; or low meat-eater – regular meat-eaters) was the response variable and mediators, and potential confounders were the predictors. In each respective mediation model, the mediators of interest were modelled as continuous variables. We then used the estimated coefficients of this model to predict the odds ratios of the binary diet group for each participant and constructed weights for binary diet groups as the inverse of these predicted odds ratios. Here the reference category (regular meat-eaters) was assigned the weight of 1 and the opposing diet group (vegetarians, fish-eaters, or low-meat eaters) were assigned the value of the inverse odds of the logistic models modelled with the mediators and adjusted confounders. We then conducted a Cox proportional hazard regression using age as the underlying time variable with all cancer site or specific cancer site, adjusting for confounders, weighting each observation by the weights derived from the logistic regression. From this Cox regression, the direct effect was obtained as the hazard ratio (HR) for the binary diet group (e.g. vegetarians – regular meat-eaters) of interest in this model and determined the indirect effect by taking total effect minus the direct effect. For the total, direct, and indirect effects, the resulting HRs were estimated and bootstrapped standard errors (300 replications) to determine 95% confidence intervals. The IORW method was conducted separately for each outcome (all cancer, prostate cancer, postmenopausal breast cancer, or colorectal cancer) and binary diet group of interest and excluded participants not included in the binary diet group of interest, and women for the prostate cancer analyses, and men for the postmenopausal breast cancer analyses. Participants with missing values for the potential mediator (i.e., BMI, IGF-I, free testosterone) were excluded from the respective mediation analyses.

### Supplementary Tables

**Supplementary Table 1.** Baseline characteristics across diet groups separated by sex

| | Male | | | | Female | | | |
|---|---|---|---|---|---|---|---|---|
| | Regular meat-eater | Low meat-eaters | Fish-eater | Vegetarian | Regular meat-eater | Low meat-eaters | Fish-eater | Vegetarian |
| Number of participants | 132,722 | 79,220 | 3,032 | 2,963 | 114,849 | 126,165 | 7,664 | 5,722 |
| Age – years, mean (SD) | 56.2 (8.2) | 57.3 (8.1) | 54.3 (8.1) | 53.1 (8.0) | 55.9 (8.1) | 56.6 (7.9) | 53.9 (8.0) | 53.0 (7.9) |
| BMI - $kg/m^2$, mean (SD) | 28.2 (4.3) | 27.5 (4.1) | 25.8 (3.6) | 25.9 (3.9) | 27.7 (5.4) | 26.7 (5.0) | 25.1 (4.5) | 25.6 (5.0) |
| Height - cm, mean (SD) | 175.7 (6.8) | 175.4 (6.9) | 176.4 (6.9) | 175.5 (7.2) | 162.4 (6.3) | 162.5 (6.3) | 163.5 (6.4) | 162.1 (6.8) |
| Physical activity | | | | | | | | |
| Low | 36801 (27.7%) | 21972 (27.7%) | 628 (20.7%) | 777 (26.2%) | 36010 (31.4%) | 37780 (29.9%) | 1802 (23.5%) | 1594 (27.9%) |
| Moderate | 62156 (46.8%) | 37698 (47.6%) | 1636 (54.0%) | 1505 (50.8%) | 54435 (47.4%) | 60994 (48.3%) | 4074 (53.2%) | 2841 (49.7%) |
| High | 28947 (21.8%) | 16599 (21.0%) | 704 (23.2%) | 610 (20.6%) | 19065 (16.6%) | 22105 (17.5%) | 1569 (20.5%) | 1080 (18.9%) |
| Townsend deprivation index | | | | | | | | |
| Q1- Most affluent | 27202 (20.5%) | 15766 (19.9%) | 458 (15.1%) | 430 (14.5%) | 23915 (20.8%) | 24667 (19.6%) | 1319 (17.2%) | 828 (14.5%) |
| Q5 - Most deprived | 26819 (20.2%) | 16416 (20.7%) | 740 (24.4%) | 827 (27.9%) | 21408 (18.6%) | 25211 (20.0%) | 1645 (21.5%) | 1389 (24.3%) |
| Ethnicity | | | | | | | | |
| White | 125463 (94.5%) | 74035 (93.5%) | 2792 (92.1%) | 2321 (78.3%) | 108496 (94.5%) | 118998 (94.3%) | 7130 (93.0%) | 4582 (80.1%) |
| Mixed or other | 1776 (1.3%) | 1112 (1.4%) | 45 (1.5%) | 45 (1.5%) | 1800 (1.6%) | 2182 (1.7%) | 127 (1.7%) | 107 (1.9%) |
| Asian or British Asian | 2422 (1.8%) | 2615 (3.3%) | 125 (4.1%) | 558 (18.8%) | 1692 (1.5%) | 2439 (1.9%) | 244 (3.2%) | 966 (16.9%) |
| Black or Black British | 2092 (1.6%) | 1101 (1.4%) | 46 (1.5%) | 12 (0.4%) | 2126 (1.9%) | 2194 (1.7%) | 121 (1.6%) | 36 (0.6%) |
| Education | | | | | | | | |
| National exam at 16 years | 18401 (13.9%) | 10397 (13.1%) | 273 (9.0%) | 323 (10.9%) | 23363 (20.3%) | 23874 (18.9%) | 907 (11.8%) | 776 (13.6%) |
| National exam at 17-18 | 6811 (5.1%) | 3807 (4.8%) | 145 (4.8%) | 188 (6.3%) | 6939 (6.0%) | 6998 (5.5%) | 433 (5.6%) | 363 (6.3%) |
| Degree or college | 83092 (62.6%) | 48766 (61.6%) | 2340 (77.2%) | 2144 (72.4%) | 63122 (55.0%) | 71025 (56.3%) | 5675 (74.0%) | 3965 (69.3%) |
| Employment | | | | | | | | |
| In paid employment | 83026 (62.6%) | 46243 (58.4%) | 2145 (70.7%) | 2146 (72.4%) | 63052 (54.9%) | 69336 (55.0%) | 5193 (67.8%) | 3919 (68.5%) |
| Retired | 38471 (29.0%) | 26228 (33.1%) | 633 (20.9%) | 514 (17.3%) | 39012 (34.0%) | 44412 (35.2%) | 1708 (22.3%) | 1068 (18.7%) |
| Not in paid employment | 9691 (7.3%) | 5887 (7.4%) | 206 (6.8%) | 274 (9.2%) | 11377 (9.9%) | 11141 (8.8%) | 671 (8.8%) | 647 (11.3%) |

| | | | | | | | | |
|---|---|---|---|---|---|---|---|---|
| Living with a partner - Yes | 102443 (77.2%) | 59313 (74.9%) | 2163 (71.3%) | 2104 (71.0%) | 67306 (58.6%) | 70915 (56.2%) | 3910 (51.0%) | 2770 (48.4%) |
| Smoking status | | | | | | | | |
| Never | 63688 (48.0%) | 39608 (50.0%) | 1658 (54.7%) | 1727 (58.3%) | 68606 (59.7%) | 74777 (59.3%) | 4417 (57.6%) | 3834 (67.0%) |
| Previous | 50320 (37.9%) | 30201 (38.1%) | 1096 (36.1%) | 951 (32.1%) | 34999 (30.5%) | 39441 (31.3%) | 2704 (35.3%) | 1529 (26.7%) |
| Light smoker <15 cigarettes/day | 3887 (2.9%) | 2048 (2.6%) | 72 (2.4%) | 79 (2.7%) | 3707 (3.2%) | 4251 (3.4%) | 218 (2.8%) | 131 (2.3%) |
| med smoker 15-29 cigarettes/day | 6093 (4.6%) | 2715 (3.4%) | 51 (1.7%) | 53 (1.8%) | 4008 (3.5%) | 3929 (3.1%) | 106 (1.4%) | 86 (1.5%) |
| heavy smoker 30+ cigarettes/day | 7738 (5.8%) | 4208 (5.3%) | 141 (4.7%) | 134 (4.5%) | 2680 (2.3%) | 3224 (2.6%) | 192 (2.5%) | 118 (2.1%) |
| Alcohol intake grams/day, Mean (SD) | 26.4 (24.7) | 21.4 (21.1) | 20.1 (18.9) | 18.5 (21.1) | 11.7 (11.9) | 10.4 (10.6) | 11.0 (11.3) | 9.9 (11.2) |
| Diabetic - Yes | 10377 (7.8%) | 5943 (7.5%) | 121 (4.0%) | 207 (7.0%) | 5226 (4.6%) | 4805 (3.8%) | 169 (2.2%) | 258 (4.5%) |
| PSA test at baseline or during follow-up - Yes | 51,555 (38.8%) | 33,394 (42.2%) | 1,125 (37.1%) | 929 (31.4%) | - | - | - | - |
| Age-adjusted proportion of PSA test at baseline or during follow-up - Yes | 39.5% | 40.5% | 41.6% | 37.2% | | | | |
| Insulin-like growth factor-I (nmol/L), mean (SD)[1] | 21.9 (5.5) | 21.9 (5.5) | 22.0 (5.7) | 21.2 (5.5) | 21.1 (5.8) | 21.0 (5.7) | 20.6 (5.8) | 19.7 (5.8) |
| Free testosterone (pmol/L), mean (SD)[1] | 220.0 (65.9) | 219.7 (63.5) | 218.7 (61.5) | 217.4 (62.3) | 15.3 (11.4) | 14.9 (10.9) | 14.0 (8.6) | 14.4 (9.4) |
| **Women specific covariates** | | | | | | | | |
| Age at first menarche | - | - | - | - | 12.6 (2.8) | 12.5 (2.9) | 12.5 (2.9) | 12.4 (3.3) |
| Menopause status at recruitment | | | | | | | | |
| Premenopausal | - | - | - | - | 24939 (21.7%) | 23360 (18.5%) | 2232 (29.1%) | 1843 (32.2%) |
| Postmenopausal | - | - | - | - | 78626 (68.5%) | 92413 (73.2%) | 4717 (61.5%) | 3342 (58.4%) |
| MHT use | | | | | | | | |
| Never | - | - | - | - | 70830 (61.7%) | 76747 (60.8%) | 5468 (71.3%) | 4386 (76.7%) |
| Former | - | - | - | - | 34149 (29.7%) | 38960 (30.9%) | 1590 (20.7%) | 966 (16.9%) |
| Current | - | - | - | - | 8993 (7.8%) | 9988 (7.9%) | 584 (7.6%) | 320 (5.6%) |
| Parity | | | | | | | | |

| | | | | | | | | |
|---|---|---|---|---|---|---|---|---|
| Nulliparous | - | - | - | - | 17671 (15.4%) | 25569 (20.3%) | 2330 (30.4%) | 1736 (30.3%) |
| 1-2 children | - | - | - | - | 67306 (58.6%) | 70915 (56.2%) | 3910 (51.0%) | 2770 (48.4%) |
| 3+ children | - | - | - | - | 29343 (25.5%) | 29526 (23.4%) | 1415 (18.5%) | 1203 (21.0%) |
| Age at first birth - years, mean (SD) | - | - | - | - | 25.4 (4.6) | 25.2 (4.6) | 26.4 (5.1) | 26.0 (4.9) |

Percentages include missing values and therefore may not add up to 100%.

Values are N (%) unless otherwise indicated.

[1] Biomarker levels are adjusted for age at recruitment, region of recruitment, height, physical activity, Townsend deprivation index, education, employment status, smoking status, alcohol consumption, ethnicity, diabetes status, menopausal hormone therapy use and menopausal status, body mass index and season of blood collection.

Abbreviations: BMI: body mass index; MHT: menopausal hormonal therapy; Q: quintile; PSA, prostate specific antigen; SD: standard deviation.

**Supplementary Table 2.** Hazard ratio and 95% confidence intervals for sequential adjustment between association of diet groups and risk of all cancer, colorectal cancer, breast cancer, and prostate cancer

| **All cancer** | **Regular meat-eaters** | **Low meat-eaters** | **Fish-eaters** | **Vegetarians** | $\chi^2$ | **P-value for heterogeneity** |
|---|---|---|---|---|---|---|
| Minimally adjusted model | 1 (ref) | 0.97 (0.95 - 0.99) | 0.87 (0.82 - 0.93) | 0.80 (0.74 - 0.86) | 56.00 | <0.001 |
| Multivariable-adjusted model | 1 (ref) | 0.98 (0.96 - 1.00) | 0.90 (0.84 - 0.96) | 0.86 (0.80 - 0.93) | 26.88 | <0.001 |
| Multivariable-adjusted model + BMI | 1 (ref) | 0.99 (0.97 - 1.01) | 0.92 (0.86 - 0.99) | 0.88 (0.82 - 0.96) | 15.53 | 0.001 |
| **Colorectal cancer** | **Regular meat-eaters** | **Low meat-eaters** | **Fish-eaters** | **Vegetarians** | $\chi^2$ | **P-value for heterogeneity** |
| Minimally adjusted model | 1 (ref) | 0.89 (0.84 - 0.94) | 0.81 (0.66 - 1.00) | 0.72 (0.56 - 0.93) | 26.12 | <0.001 |
| Multivariable-adjusted model | 1 (ref) | 0.91 (0.86 - 0.96) | 0.84 (0.68 - 1.03) | 0.78 (0.61 - 1.01) | 16.80 | 0.001 |
| Multivariable-adjusted model + BMI | 1 (ref) | 0.92 (0.87 - 0.97) | 0.86 (0.70 - 1.06) | 0.81 (0.63 - 1.04) | 12.69 | 0.005 |
| **Postmenopausal breast cancer** | **Regular meat-eaters** | **Low meat-eaters** | **Fish-eaters** | **Vegetarians** | $\chi^2$ | **P-value for heterogeneity** |
| Minimally adjusted model | 1 (ref) | 0.97 (0.92 - 1.01) | 0.92 (0.80 - 1.07) | 0.80 (0.67 - 0.97) | 7.43 | 0.059 |
| Multivariable-adjusted model | 1 (ref) | 0.96 (0.92 - 1.01) | 0.92 (0.79 - 1.06) | 0.82 (0.68 - 0.99) | 6.65 | 0.084 |
| Multivariable-adjusted model + BMI | 1 (ref) | 0.99 (0.94 - 1.03) | 0.97 (0.84 - 1.12) | 0.87 (0.72 - 1.05) | 2.46 | 0.483 |
| **Prostate cancer** | **Regular meat-eaters** | **Low meat-eaters** | **Fish-eaters** | **Vegetarians** | $\chi^2$ | **P-value for heterogeneity** |
| Minimally adjusted model | 1 (ref) | 1.00 (0.96 - 1.04) | 0.80 (0.65 - 0.98) | 0.61 (0.48 - 0.79) | 22.54 | <0.001 |
| Multivariable-adjusted model | 1 (ref) | 1.00 (0.96 - 1.05) | 0.80 (0.65 - 0.99) | 0.69 (0.54 - 0.89) | 14.08 | 0.003 |
| Multivariable-adjusted model + BMI | 1 (ref) | 1.00 (0.96 - 1.04) | 0.80 (0.65 - 0.98) | 0.69 (0.54 - 0.89) | 14.28 | 0.003 |
| Multivariable-adjusted + PSA test | 1 (ref) | 1.00 (0.96 -1.04) | 0.80 (0.65 – 0.98) | 0.70 (0.55-0.90) | 13.47 | 0.004 |

Regular meat-eaters: consumed red or processed meat or poultry >5 times a week. Low meat-eaters: consumed red and processed meat or poultry ≤5 times per week. Fish-eaters: do not consume red, processed meat, or poultry but consumed fish. Vegetarians (including vegans): do not consume any meat or fish.

Minimally adjusted model used age as the underlying time variable and are stratified by sex (for all cancer and colorectal cancer), age groups, and controlled for region of recruitment. Models are restricted to only men for prostate cancer and only women for postmenopausal breast cancer.

Multivariable-adjusted models for all cancer outcomes: Minimally adjusted model + height, physical activity, Townsend deprivation index, education, employment status, smoking status, alcohol consumption, ethnicity, and diabetes status. Full details for each covariate are provided in the statistical analysis section in the main text.

For all cancer and colorectal cancer multivariable-adjusted model additionally adjust for menopausal hormone therapy (MHT) use and menopausal status. For colorectal cancer, multi-variable-adjusted was also adjusted for NSAID use.

For prostate cancer multivariable-adjusted models additionally adjust for marital status.

For breast cancer multivariable-adjusted model additionally adjust for MHT use, age at menarche, and parity.

Multivariable-adjusted model + BMI adds body mass index (<20, 20-22.49,22.5-24.9, 25.0-27.49, 27.5-29.9, 30-32.49, 32.5-34.9, ≥35 $kg/m^2$ or unknown) to multivariable models.

Multivariable-adjusted model + PSA test adds prostate specific antigen testing reported at baseline and from follow-up general practice records available in a subsample of men.

$\chi^2$ (df=3) and p-values from likelihood ratio tests for model fit comparing a model without diet groups, to a model including diet groups.

Abbreviations: BMI: body mass index; df: degrees of freedom; MHT: menopausal hormonal therapy; NSAID, non-steroid anti-inflammatory drugs; PSA, prostate specific antigen test; ref, reference group.

**Supplementary Table 3.** Subgroup analyses for diet groups on risk of **all cancers**

| | BMI | | | | | | | |
|---|---|---|---|---|---|---|---|---|
| | <27.5 kg/m$^2$ | | | ≥27.5 kg/m$^2$ | | | | |
| | **N** | **Cases** | **HR (95% CI)** | **N** | **Cases** | **HR (95% CI)** | $\chi^2$ | **P-value** |
| Regular meat-eaters | 127785 | 14174 | 1 (ref) | 118297 | 15055 | 1 (ref) | 1.07 | 0.785 |
| Low meat-eaters | 123610 | 13585 | 0.99 (0.97 - 1.02) | 80720 | 10244 | 0.98 (0.95 - 1.00) | | |
| Fish-eaters | 8001 | 690 | 0.94 (0.87 - 1.01) | 2644 | 241 | 0.85 (0.75 - 0.97) | | |
| Vegetarians | 6152 | 459 | 0.87 (0.80 - 0.96) | 2475 | 208 | 0.91 (0.79 - 1.04) | | |
| | **Smoking status** | | | | | | | |
| | **Never** | | | **Ever** | | | | |
| | **N** | **Cases** | **HR (95% CI)** | **N** | **Cases** | **HR (95% CI)** | $\chi^2$ | **P-value** |
| Regular meat-eaters | 132294 | 13154 | 1 (ref) | 113432 | 15995 | 1 (ref) | 7.55 | 0.056 |
| Low meat-eaters | 114385 | 11699 | 1.01 (0.98 - 1.03) | 90017 | 12102 | 0.97 (0.94 - 0.99) | | |
| Fish-eaters | 6075 | 498 | 0.95 (0.87 - 1.04) | 4580 | 439 | 0.86 (0.78 - 0.95) | | |
| Vegetarians | 5561 | 405 | 0.94 (0.85 - 1.04) | 3081 | 262 | 0.79 (0.70 - 0.90) | | |
| | **Smoking status – censoring lung cancer cases** | | | | | | | |
| | **Never** | | | **Ever** | | | | |
| | **N** | **Cases** | **HR (95% CI)** | **N** | **Cases** | **HR (95% CI)** | $\chi^2$ | **P-value** |
| Regular meat-eaters | 132050 | 12911 | 1 (ref) | 111542 | 14110 | 1 (ref) | 4.38 | 0.223 |
| Low meat-eaters | 114105 | 11419 | 1.00 (0.98 - 1.03) | 88648 | 10735 | 0.98 (0.96 - 1.01) | | |
| Fish-eaters | 6062 | 485 | 0.95 (0.87 - 1.04) | 4536 | 395 | 0.89 (0.80 - 0.98) | | |
| Vegetarians | 5550 | 394 | 0.93 (0.84 - 1.03) | 3057 | 238 | 0.82 (0.72 - 0.93) | | |

All models used age as the underlying time variable and are stratified by sex and age groups, and adjusted for region of recruitment, height, physical activity, Townsend deprivation index, education, employment status, smoking status (except when smoking status was the subgroup of interest), alcohol consumption, ethnicity, diabetes status, menopausal hormone therapy use, and menopausal status. Full details for each covariate are provided in the statistical analysis section in the main text.

$\chi^2$ and p-values from likelihood ratio tests for model fit comparing a model without an interaction term between subgroup and diet groups, to a model including an interaction between subgroup and diet groups.

Abbreviations: BMI: body mass index; CI: confidence intervals; HR: hazard ratios; ref: reference group.

**Supplementary Table 4.** Subgroup analyses for diet groups on risk of **colorectal cancer**

| | Sex | | | | | | | |
|---|---|---|---|---|---|---|---|---|
| | Female | | | Male | | | | |
| | **N** | **Cases** | **HR (95% CI)** | **N** | **Cases** | **HR (95% CI)** | $\chi^2$ | **P-value** |
| Regular meat-eaters | 114849 | 1168 | 1 (ref) | 132722 | 2129 | 1 (ref) | 12.17 | 0.007 |
| Low meat-eaters | 126165 | 1280 | 0.96 (0.89 - 1.04) | 79220 | 1150 | 0.89 (0.83 - 0.95) | | |
| Fish-eaters | 7664 | 66 | 0.97 (0.75 - 1.24) | 3032 | 26 | 0.69 (0.47 - 1.01) | | |
| Vegetarians | 5722 | 45 | 0.97 (0.72 - 1.31) | 2963 | 18 | 0.57 (0.36 - 0.91) | | |
| | **BMI** | | | | | | | |
| | **<27.5 kg/m²** | | | **≥27.5 kg/m²** | | | | |
| | **N** | **Cases** | **HR (95% CI)** | **N** | **Cases** | **HR (95% CI)** | $\chi^2$ | **P-value** |
| Regular meat-eaters | 127785 | 1553 | 1 (ref) | 118297 | 1728 | 1 (ref) | 2.79 | 0.425 |
| Low meat-eaters | 123610 | 1358 | 0.90 (0.84 - 0.97) | 80720 | 1058 | 0.92 (0.86 - 1.00) | | |
| Fish-eaters | 8001 | 69 | 0.87 (0.68 - 1.11) | 2644 | 20 | 0.69 (0.44 - 1.07) | | |
| Vegetarians | 6152 | 48 | 0.86 (0.64 - 1.15) | 2475 | 15 | 0.65 (0.39 - 1.09) | | |
| | **Smoking status** | | | | | | | |
| | **Never** | | | **Ever** | | | | |
| | **N** | **Cases** | **HR (95% CI)** | **N** | **Cases** | **HR (95% CI)** | $\chi^2$ | **P-value** |
| Regular meat-eaters | 132294 | 1472 | 1 (ref) | 113432 | 1799 | 1 (ref) | 2.17 | 0.538 |
| Low meat-eaters | 114385 | 1180 | 0.92 (0.85 - 0.99) | 90017 | 1243 | 0.92 (0.86 - 1.00) | | |
| Fish-eaters | 6075 | 51 | 0.91 (0.69 - 1.21) | 4580 | 41 | 0.82 (0.60 - 1.11) | | |
| Vegetarians | 5561 | 31 | 0.68 (0.47 - 0.97) | 3081 | 32 | 0.99 (0.70 - 1.41) | | |

All models used age as the underlying time variable and are stratified by sex (except when sex was the subgroup of interest) and age groups, and adjusted for region of recruitment, height, physical activity, Townsend deprivation index, education, employment status, smoking status (except when smoking was subgroup of interest), alcohol consumption, ethnicity, diabetes status, menopausal hormone therapy use, menopausal status, non-steroid anti-inflammatory drug use. Full details for each covariate are provided in the statistical analysis section in the main text.

$\chi^2$ and p-values from likelihood ratio tests for model fit comparing a model without an interaction term between subgroup and diet groups, to a model including an interaction between subgroup and diet groups.

Abbreviations: BMI: body mass index; CI: confidence intervals; HR: hazard ratios; ref: reference group.

**Supplementary Table 5**. Subgroup analyses for diet groups on risk of **postmenopausal breast cancer** in women

| | **BMI** | | | | | | | |
|---|---|---|---|---|---|---|---|---|
| | **<27.5 kg/m$^2$** | | | **≥27.5 kg/m$^2$** | | | | |
| | **N** | **Cases** | **HR (95% CI)** | **N** | **Cases** | **HR (95% CI)** | **$\chi^2$** | **P-value** |
| Regular meat-eaters | 59198 | 1735 | 1 (ref) | 47360 | 1680 | 1 (ref) | 5.23 | 0.156 |
| Low meat-eaters | 75163 | 2198 | 0.96 (0.90 - 1.02) | 43399 | 1583 | 1.00 (0.94 - 1.08) | | |
| Fish-eaters | 5201 | 148 | 1.03 (0.87 - 1.22) | 1663 | 42 | 0.75 (0.55 - 1.01) | | |
| Vegetarians | 3560 | 79 | 0.85 (0.68 - 1.07) | 1456 | 37 | 0.86 (0.61 - 1.20) | | |

| | **Smoking status** | | | | | | | |
|---|---|---|---|---|---|---|---|---|
| | **Never** | | | **Ever** | | | | |
| | **N** | **Cases** | **HR (95% CI)** | **N** | **Cases** | **HR (95% CI)** | **$\chi^2$** | **P-value** |
| Regular meat-eaters | 63753 | 1909 | 1 (ref) | 42559 | 1501 | 1 (ref) | 1.32 | 0.725 |
| Low meat-eaters | 70440 | 2159 | 0.98 (0.93 - 1.05) | 48173 | 1631 | 0.94 (0.88 - 1.01) | | |
| Fish-eaters | 3993 | 104 | 0.93 (0.77 - 1.14) | 2879 | 86 | 0.90 (0.72 - 1.12) | | |
| Vegetarians | 3401 | 76 | 0.86 (0.68 - 1.09) | 1625 | 39 | 0.75 (0.55 - 1.04) | | |

All models used age as the underlying time variable and are stratified, age groups, and adjusted for region of recruitment, height, physical activity, Townsend deprivation index, education, employment status, smoking status (except when smoking status was the subgroup of interest), alcohol consumption, ethnicity, diabetes status, menopausal hormone therapy use, age at menarche, and age at first birth/parity. Full details for each covariate are provided in the statistical analysis section in the main text.

Models are restricted to only women.

$\chi^2$ and p-values from likelihood ratio tests for model fit comparing a model without an interaction term between subgroup and diet groups, to a model including an interaction between subgroup and diet groups.

Abbreviations: BMI: body mass index; CI: confidence intervals; HR: hazard ratios; ref: reference group.

**Supplementary Table 6.** Subgroup analyses across diet groups on risk of **prostate cancer** in men

| | BMI | | | | | | | |
|---|---|---|---|---|---|---|---|---|
| | <27.5 kg/m$^2$ | | | ≥27.5 kg/m$^2$ | | | | |
| | **N** | **Cases** | **HR (95% CI)** | **N** | **Cases** | **HR (95% CI)** | $\chi^2$ | **P-value** |
| Regular meat-eaters | 63771 | 2750 | 1 (ref) | 68053 | 2873 | 1 (ref) | 4.02 | 0.260 |
| Low meat-eaters | 43570 | 2098 | 1.02 (0.96 - 1.08) | 35201 | 1583 | 0.98 (0.92 - 1.05) | | |
| Fish-eaters | 2174 | 60 | 0.70 (0.54 - 0.91) | 844 | 32 | 1.04 (0.73 - 1.48) | | |
| Vegetarians | 2087 | 49 | 0.72 (0.54 - 0.95) | 858 | 15 | 0.62 (0.37 - 1.04) | | |
| | **Smoking status** | | | | | | | |
| | **Never** | | | **Ever** | | | | |
| | **N** | **Cases** | **HR (95% CI)** | **N** | **Cases** | **HR (95% CI)** | $\chi^2$ | **P-value** |
| Regular meat-eaters | 63688 | 2593 | 1 (ref) | 68038 | 3009 | 1 (ref) | 0.99 | 0.804 |
| Low meat-eaters | 39608 | 1809 | 1.00 (0.95 - 1.07) | 39172 | 1861 | 1.00 (0.94 - 1.06) | | |
| Fish-eaters | 1658 | 53 | 0.83 (0.63 - 1.09) | 1360 | 40 | 0.79 (0.58 - 1.08) | | |
| Vegetarians | 1727 | 40 | 0.76 (0.56 - 1.05) | 1217 | 24 | 0.61 (0.41 - 0.92) | | |

All models used age as the underlying time variable and are stratified by age groups, and adjusted for region of recruitment, height, physical activity, Townsend deprivation index, education, employment status, smoking status (except when smoking status was the subgroup of interest), alcohol consumption, ethnicity, diabetes status, and marital status. Full details for each covariate are provided in the statistical analysis section in the main text.

Models are restricted to only men.

$\chi^2$ and p-values from likelihood ratio tests for model fit comparing a model without an interaction term between subgroup and diet groups, to a model including an interaction between subgroup and diet groups.

Abbreviations: BMI: body mass index; CI: confidence intervals; HR: hazard ratios; ref: reference group.

**Supplementary Table 7.** Adjusted and relative means (95% CI) of BMI, IGF-I, and free testosterone concentrations measured at recruitment across diet groups

| | **BMI (kg/m$^2$)** | | **IGF-I (nmol/L)** | | **Free testosterone (pmol/L)** | |
|---|---|---|---|---|---|---|
| **All** | Adjusted mean | Relative Mean | Adjusted mean | Relative Mean | Adjusted mean | Relative Mean |
| Regular meat-eater | 27.90 (27.88-27.92) | 1 (ref) | 21.49 (21.47-21.51) | 1 (ref) | 119.67 (119.47-119.87) | 1 (ref) |
| Low meat-eater | 27.03 (27.01-27.05) | 0.97 (0.97 - 0.97) | 21.43 (21.40-21.45) | 1.00 (1.00 - 1.00) | 119.19 (118.97-119.42) | 1.00 (0.99 - 1.00) |
| Fish-eater | 25.68 (25.60-25.77) | 0.92 (0.92 - 0.92) | 21.20 (21.10-21.31) | 0.99 (0.98 - 0.99) | 117.84 (116.85-118.82) | 0.98 (0.98 - 0.99) |
| Vegetarian | 25.87 (25.77-25.97) | 0.93 (0.92 - 0.93) | 20.29 (20.17-20.41) | 0.94 (0.94 - 0.95) | 117.69 (116.58-118.80) | 0.98 (0.97 - 0.99) |
| **Females** | | | | | | |
| Regular meat-eater | 27.71 (27.68-27.74) | 1 (ref) | 21.16 (21.12-21.19) | 1 (ref) | 15.28 (15.21-15.35) | 1 (ref) |
| Low meat-eater | 26.69 (26.66-26.72) | 0.96 (0.96 – 0.96) | 21.00 (20.97-21.03) | 0.99 (0.99 - 0.99) | 14.88 (14.81-14.95) | 0.97 (0.97 - 0.98) |
| Fish-eater | 25.43 (25.32-25.54) | 0.92 (0.91 - 0.92) | 20.66 (20.53-20.78) | 0.98 (0.97 - 0.98) | 13.98 (13.70-14.27) | 0.92 (0.90 - 0.93) |
| Vegetarian | 25.61 (25.48-25.74) | 0.92 (0.91 - 0.93) | 19.69 (19.54-19.84) | 0.93 (0.92 - 0.94) | 14.39 (14.05-14.73) | 0.94 (0.92 - 0.96) |
| **Males** | | | | | | |
| Regular meat-eater | 28.14 (28.12-28.17) | 1 (ref) | 21.91 (21.88-21.94) | 1 (ref) | 220.00 (219.65-220.36) | 1 (ref) |
| Low meat-eater | 27.47 (27.45-27.50) | 0.98 (0.98 - 0.98) | 21.92 (21.88-21.96) | 1.00 (1.00 - 1.00) | 219.77 (219.31-220.23) | 1.00 (1.00 - 1.00) |
| Fish-eater | 26.05 (25.91-26.20) | 0.93 (0.93 - 0.93) | 22.01 (21.81-22.20) | 1.00 (1.00 - 1.01) | 218.65 (216.32-220.99) | 0.99 (0.98 - 1.00) |
| Vegetarian | 26.10 (25.95-26.25) | 0.93 (0.92 - 0.93) | 21.17 (20.97-21.37) | 0.97 (0.96 - 0.98) | 217.41 (215.01-219.81) | 0.99 (0.98 – 1.00) |

All biomarkers are adjusted for: age groups, sex (for all participants), region of recruitment, height, physical activity, Townsend deprivation index, education, employment status, smoking status, alcohol consumption, ethnicity, diabetes status, menopausal hormone therapy (except for males) use and menopausal status (except for males), and body mass index (except when BMI was the outcome).

Abbreviations: BMI, body mass index; CI, confidence intervals; IGF-I, insulin-like growth factor-I; ref, reference group.

### Supplementary Figures

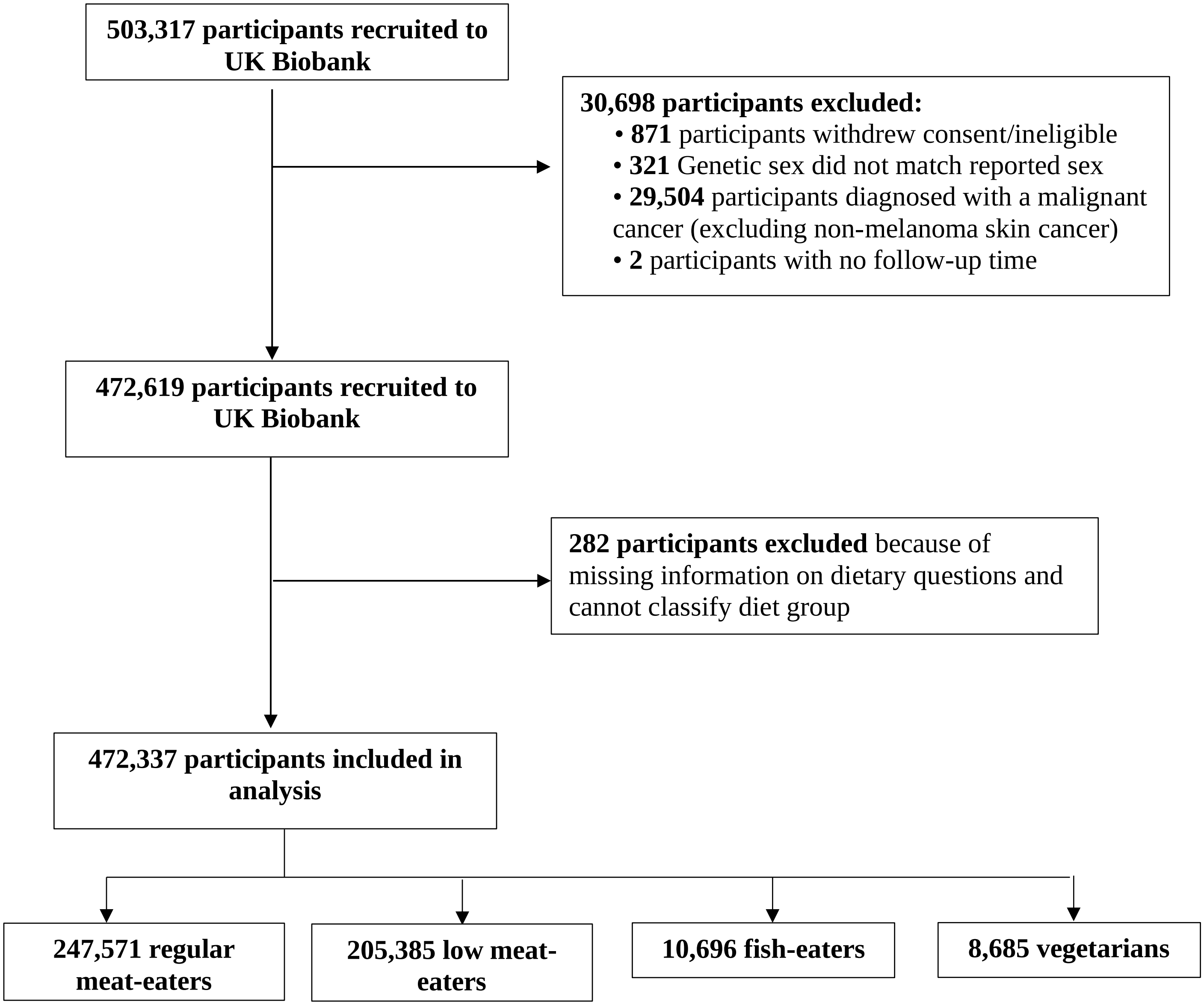

**Supplementary Figure 1.** Flow diagram of the study exclusion criteria.

**Only white ethnicity**

| | N | Cases | HR (95% CI) | $c^2_3$ | p-values |
|---|---|---|---|---|---|
| **All cancer** | | | | | |
| Regular meat-eaters | 233,959 | 28304 | 1 (ref) | 28.01 | <0.001 |
| Low meat-eaters | 193,033 | 22942 | 0.98 (0.96 - 1.00) | | |
| Fish-eaters | 9,922 | 879 | 0.90 (0.84 - 0.96) | | |
| Vegetarians | 6,903 | 544 | 0.85 (0.78 - 0.92) | | |
| **Colorectal cancer** | | | | | |
| Regular meat-eaters | 233,959 | 3186 | 1 (ref) | 10.5 | 0.015 |
| Low meat-eaters | 193,033 | 2342 | 0.92 (0.87 - 0.97) | | |
| Fish-eaters | 9,922 | 85 | 0.85 (0.68 - 1.05) | | |
| Vegetarians | 6,903 | 57 | 0.87 (0.67 - 1.14) | | |
| **Postmenopausal breast cancer** | | | | | |
| Regular meat-eaters | 101,614 | 3312 | 1 (ref) | 8.19 | 0.042 |
| Low meat-eaters | 112,767 | 3668 | 0.97 (0.92 - 1.02) | | |
| Fish-eaters | 6,429 | 176 | 0.90 (0.77 - 1.05) | | |
| Vegetarians | 4,026 | 89 | 0.78 (0.63 - 0.96) | | |
| **Prostate cancer** | | | | | |
| Regular meat-eaters | 125,463 | 5401 | 1 (ref) | 12.56 | 0.006 |
| Low meat-eaters | 74,035 | 3506 | 1.00 (0.96 - 1.04) | | |
| Fish-eaters | 2,792 | 86 | 0.81 (0.65 - 1.00) | | |
| Vegetarians | 2,321 | 52 | 0.68 (0.52 - 0.89) | | |

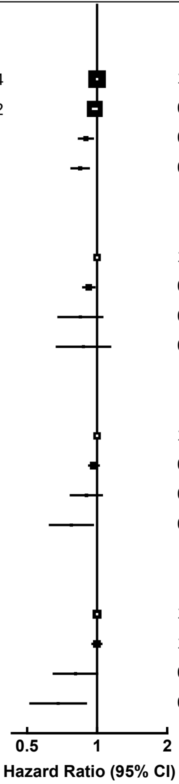

**Complete case analyses**

| | N | Cases | HR (95% CI) | $c^2_3$ | p-values |
|---|---|---|---|---|---|
| **All cancer** | | | | | |
| Regular meat-eaters | 192,450 | 21227 | 1 (ref) | 15.83 | 0.001 |
| Low meat-eaters | 156,887 | 17215 | 1.00 (0.98 - 1.02) | | |
| Fish-eaters | 9,326 | 787 | 0.92 (0.86 - 0.99) | | |
| Vegetarians | 7,326 | 535 | 0.86 (0.79 - 0.94) | | |
| **Colorectal cancer** | | | | | |
| Regular meat-eaters | 192,450 | 2427 | 1 (ref) | 9.5 | 0.023 |
| Low meat-eaters | 156,887 | 1755 | 0.91 (0.86 - 0.97) | | |
| Fish-eaters | 9,326 | 76 | 0.86 (0.68 - 1.08) | | |
| Vegetarians | 7,326 | 54 | 0.84 (0.64 - 1.11) | | |
| **Postmenopausal breast cancer** | | | | | |
| Regular meat-eaters | 63,372 | 1971 | 1 (ref) | 5.01 | 0.171 |
| Low meat-eaters | 70,502 | 2174 | 0.96 (0.90 - 1.02) | | |
| Fish-eaters | 4,671 | 128 | 0.93 (0.77 - 1.11) | | |
| Vegetarians | 3,228 | 78 | 0.87 (0.69 - 1.09) | | |
| **Prostate cancer** | | | | | |
| Regular meat-eaters | 103,853 | 4251 | 1 (ref) | 14.16 | 0.003 |
| Low meat-eaters | 60,131 | 2788 | 1.02 (0.97 - 1.07) | | |
| Fish-eaters | 2,638 | 78 | 0.78 (0.62 - 0.98) | | |
| Vegetarians | 2,513 | 55 | 0.70 (0.53 - 0.91) | | |

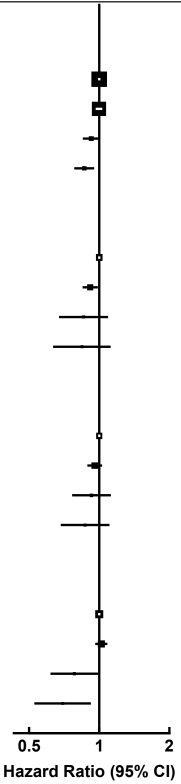

**Supplementary Figure 2.** Hazard ratios and 95% confidence intervals for sensitivity analyses including only participants of **white ethnicity** and **complete cases analyses** on associations between diet groups and risk of all cancer, prostate, postmenopausal breast, or colorectal cancer.

All models used age as the underlying time variable and are stratified by sex (for only all cancer and colorectal cancer) and age groups, and adjusted for region of recruitment, height, physical activity, Townsend deprivation index, education, employment status, smoking status, alcohol consumption, ethnicity, and diabetes status. Full details for each covariate are provided in the statistical analysis section in the main text.

For all cancer and colorectal cancer models are further adjusted for: menopausal hormone therapy use, and menopausal status. For colorectal cancer, models were also adjusted for NSAID use.

For prostate cancer models are further adjusted for: marital status.

For postmenopausal breast cancer models are further adjusted for: menopausal hormone therapy use, age at menarche, and age at first birth/parity.

Models are restricted to only men for prostate cancer and only women for postmenopausal breast cancer.

$\chi^2$ (degrees of freedom in subscript) and p-values from likelihood ratio tests for model fit comparing a model without diet groups, to a model including diet groups.

Abbreviations: CI: confidence intervals; HR: hazard ratios; N: number of participants; NSAID: non-steroid anti-inflammatory drug; ref: reference group.

**Removing first two years of follow-up**

| | N | Cases | HR (95% CI) | $c^2_3$ | p-values |
|---|---|---|---|---|---|
| **All cancer** | | | | | |
| Regular meat-eaters | 243,364 | 25726 | 1 (ref) | 27.4 | <0.001 |
| Low meat-eaters | 201,930 | 20919 | 0.98 (0.96 - 1.00) | | |
| Fish-eaters | 10,541 | 794 | 0.87 (0.81 - 0.93) | | |
| Vegetarians | 8,603 | 592 | 0.87 (0.80 - 0.95) | | |
| **Colorectal cancer** | | | | | |
| Regular meat-eaters | 243,364 | 2856 | 1 (ref) | 14.91 | 0.002 |
| Low meat-eaters | 201,930 | 2091 | 0.91 (0.86 - 0.96) | | |
| Fish-eaters | 10,541 | 75 | 0.80 (0.63 - 1.00) | | |
| Vegetarians | 8,603 | 55 | 0.80 (0.61 - 1.05) | | |
| **Postmenopausal breast cancer** | | | | | |
| Regular meat-eaters | 105,752 | 2919 | 1 (ref) | 5.63 | 0.131 |
| Low meat-eaters | 117,559 | 3265 | 0.97 (0.93 - 1.02) | | |
| Fish-eaters | 6,819 | 165 | 0.92 (0.79 - 1.08) | | |
| Vegetarians | 5,007 | 99 | 0.81 (0.66 - 0.99) | | |
| **Prostate cancer** | | | | | |
| Regular meat-eaters | 130,165 | 4919 | 1 (ref) | 16.15 | 0.001 |
| Low meat-eaters | 77,606 | 3191 | 0.99 (0.95 - 1.04) | | |
| Fish-eaters | 2,978 | 71 | 0.69 (0.55 - 0.88) | | |
| Vegetarians | 2,940 | 61 | 0.74 (0.57 - 0.95) | | |

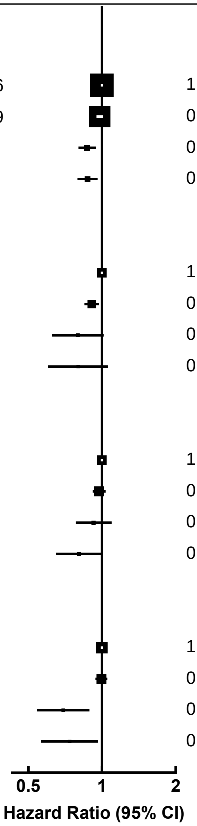

**Adding fruit and vegetable intake into model**

| | N | Cases | HR (95% CI) | $c^2_3$ | p-values |
|---|---|---|---|---|---|
| **All cancer** | | | | | |
| Regular meat-eaters | 247,571 | 29398 | 1 (ref) | 23.63 | <0.001 |
| Low meat-eaters | 205,385 | 23954 | 0.98 (0.96 - 1.00) | | |
| Fish-eaters | 10,696 | 940 | 0.90 (0.85 - 0.97) | | |
| Vegetarians | 8,685 | 669 | 0.87 (0.80 - 0.94) | | |
| **Colorectal cancer** | | | | | |
| Regular meat-eaters | 247,571 | 3297 | 1 (ref) | 12.42 | 0.006 |
| Low meat-eaters | 205,385 | 2430 | 0.92 (0.87 - 0.97) | | |
| Fish-eaters | 10,696 | 92 | 0.86 (0.70 - 1.07) | | |
| Vegetarians | 8,685 | 63 | 0.81 (0.63 - 1.04) | | |
| **Postmenopausal breast cancer** | | | | | |
| Regular meat-eaters | 107,097 | 3429 | 1 (ref) | 5.5 | 0.138 |
| Low meat-eaters | 119,134 | 3802 | 0.97 (0.92 - 1.01) | | |
| Fish-eaters | 6,897 | 190 | 0.93 (0.80 - 1.07) | | |
| Vegetarians | 5,050 | 116 | 0.83 (0.69 - 1.01) | | |
| **Prostate cancer** | | | | | |
| Regular meat-eaters | 132,722 | 5655 | 1 (ref) | 13.91 | 0.003 |
| Low meat-eaters | 79,220 | 3689 | 1.00 (0.96 - 1.04) | | |
| Fish-eaters | 3,032 | 93 | 0.80 (0.65 - 0.99) | | |
| Vegetarians | 2,963 | 64 | 0.69 (0.54 - 0.89) | | |

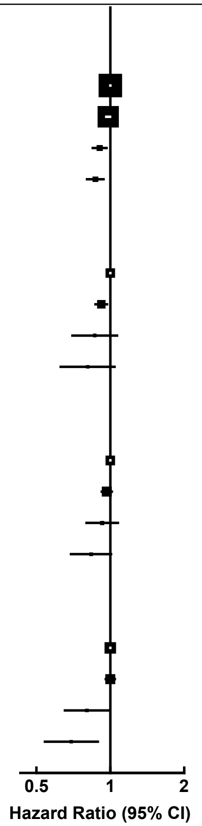

**Supplementary Figure 3.** Hazard ratios and 95% confidence intervals for sensitivity analyses **removing first two years of follow-up** and **adjusting for fruit and vegetable** intake on associations between diet groups and risk of all cancer, prostate, postmenopausal breast, or colorectal cancer.

All models used age as the underlying time variable and are stratified by sex (for only all cancer and colorectal cancer), age groups, and adjusted for region of recruitment, height, physical activity, Townsend deprivation index, education, employment status, smoking status, alcohol consumption, ethnicity, and diabetes status. Full details for each covariate are provided in the statistical analysis section in the main text.

For all cancer and colorectal cancer models are further adjusted for: menopausal hormone therapy use and menopausal status. For colorectal cancer, models were also adjusted for NSAID use.

For prostate cancer models are further adjusted for: marital status.

For postmenopausal breast cancer model are further adjusted for: menopausal hormone therapy (MHT) use, age at menarche, and age at first birth/parity.

Models are restricted to only men for prostate cancer and only women for postmenopausal breast cancer.

$\chi^2$ (degrees of freedom in subscript) and p-values from likelihood ratio tests for model fit comparing a model without diet groups, to a model including diet groups.

Abbreviations: CI: confidence intervals; HR: hazard ratios; N: number of participants; NSAID: non-steroid anti-inflammatory drug; ref: reference group.
